# A Systematic Review of the Methodological Quality of Economic Evaluations in Genetic Screening and Testing for Monogenic Disorders

**DOI:** 10.1101/2021.10.14.21265011

**Authors:** Karl Johnson, Kate Saylor, Isabella Guynn, Karen Hicklin, Jonathan S. Berg, Kristen Hassmiller Lich

**Affiliations:** University of North Carolina, Gillings School of Global Public Health, Department of Health Policy and Management; University of North Carolina, Department of Public Policy; University of North Carolina, School of Medicine, Department of Genetics

**Author notes:** Corresponding Author: Kristen Hassmiller Lich, T, F, 1105E McGavran-Greenberg Hall, CB #7411, Chapel Hill, NC 27599-7411.

## Abstract

**Purpose:** Understanding the value of genetic screening and testing for monogenic disorders requires high-quality, methodologically robust economic evaluations. This systematic review sought to assess the methodological quality among such studies and examine opportunities for improvement.

**Methods:** We searched Pubmed, Cochrane, Embase, and Web of Science for economic evaluations of genetic screening/testing (2013-2019). Methodological rigor and adherence to best practices were systematically assessed using the BMJ checklist.

**Results:** Across 47 identified studies, there was substantial variation in modeling approaches, reporting detail, and sophistication. Models ranged from simple decision trees to individual-level microsimulation, comparing between two and >20 alternative interventions. Many studies failed to report sufficient detail to enable replication or did not justify modeling assumptions, especially for costing methods and utility values. Meta-analyses, systematic reviews, or calibration were rarely used to derive parameter estimates. Nearly all studies conducted some sensitivity analysis, and more sophisticated studies implemented probabilistic sensitivity/uncertainty analysis, threshold analysis, and value of information analysis.

**Conclusion:** We describe a heterogeneous body of work and present recommendations and exemplar studies across the methodological domains of (1) perspective, scope, and parameter selection, (2) use of uncertainty/sensitivity analyses, and (3) reporting transparency for improvement in the economic evaluation of genetic screening/testing.

## INTRODUCTION

Genetic screening and testing for monogenic diseases can be used to establish a definitive molecular diagnosis in symptomatic patients, identify increased risk of disease in pre-symptomatic individuals, provide information about prognosis or management of rare disorders, identify other at-risk family members, and guide reproductive planning. If used appropriately, such analysis has the potential to reduce morbidity and mortality through disease prevention or early intervention, targeted treatment, and avoidance of inappropriate or ineffective treatment. However, genetic analysis and indicated downstream care for people who test positive can be costly for both the health system and the patient. Despite being rare (the most common affecting less than 1% of the population), molecular diagnosis of monogenic conditions can be highly useful from a clinical perspective. Currently, diagnostic genetic testing is recommended only to those meeting specific clinical criteria or after other clinical tests have failed to give a definite diagnosis. It may be cost-effective to identify and care for patients with monogenic conditions before symptoms manifest, especially for conditions with effective interventions that could improve clinical outcomes. Researchers are assessing the value of screening for clinically useful monogenic conditions within a broader population. Economic evaluations—including cost-consequence, cost-benefit, cost-utility, and cost-effectiveness analyses^1^—are critical for assessing the potential value of genetic screening/testing for specific applications. Over the last two decades, the number of such evaluations has increased rapidly.^2, 3^ Yet, the speed with which economic evaluations have been produced may be outpacing the field’s ability to disseminate and widely adopt best practices, as well as identify gaps where best practices have not been adopted.

High-quality methodological approaches to economic evaluations are essential for the appropriate interpretation and implementation of study findings. Despite the recent publication of several methodological recommendations for cost-effectiveness analyses in genetic medicine, study quality across disease areas has not been systematically reviewed.^4–6^ Importantly, there are methodological challenges unique to economic evaluations of clinical genetic screening and testing for monogenic disorders that deserve specific attention.^7^ Compared with other medical interventions that have more routinely been subjects of economic evaluations (e.g., pharmacoeconomics), the methodological tendencies of economic evaluators of genetic screening and testing programs may still be in formation.

This qualitative systematic review characterizes the methodological quality of recent economic evaluations of genetic screening and testing for monogenic disorders, spanning from birth to diagnosis. Throughout this review, we use the term “genetic testing” when referring to a clinical diagnostic setting in which a patient is at increased risk for a genetic disorder due to their personal and/or family history; we use “genetic screening” when the individual being screened is not known to be symptomatic of, nor at substantially increased risk for, such a condition. We emphasized this distinction given both the differing resources demanded of and health outcomes that may be experienced through either strategy and the field’s interest in evaluating screening programs. See **Appendix 1** for more detail. The goal of this review is to improve the methodological quality of future economic evaluations to guide implementation of such studies. Where consistent methodological limitations were identified, we have provided recommendations and exemplar models.

## METHODS

### Search Strategy

This systematic review identified economic evaluations of genetic screening and testing for monogenic disorders, focusing on assays that seek to establish (or refine) a genetic risk or diagnosis. Included studies incorporated costs and health outcomes downstream from genetic testing and diagnosis. The review was registered with PROSPERO on July 2, 2019 (record number CRD42019141086). Studies that did not include complete economic evaluations (“the comparative analysis of alternative courses of action in terms of both their costs and consequences”^1^) or considered no health outcomes beyond diagnostic yield were excluded.^8, 9^ Studies of common variants and polygenic risk scores for complex diseases were excluded, as were studies of somatic variants or gene expression in tumors.^10, 11^ Pharmacogenetic screening was excluded, defined as testing for genetic variants primarily related to adverse reactions to drugs or drug metabolism.^12^ Genetic testing/screening specifically related to reproductive planning (pre-conception or pre-natal) was excluded.^13^ Systematic reviews and commentaries were also excluded.^14^ Additional search strategy details are included in **Appendix 2**.

### Code Development

Qualitative codes reflecting methodological features of evaluations were developed using a top-down and inductive approach. Initial codes were adopted from the 1996 checklist developed by Drummond and Jefferson for the BMJ (hereafter: “BMJ checklist”), along with features highlighted in similar systematic reviews.^15–18^ While more recent checklists have been developed as guides for authors,^19^ the BMJ checklist was chosen given its emphasis on quality assessment by reviewers, its use in recent reviews of genetic evaluations^18, 20^ and its widespread use among similar systematic reviews published from 2010 to 2018.^21^ A full list of the codes and summary statement templates used can be found in **Appendix 3**.

### BMJ Checklist and Qualitative Assessment beyond the BMJ Checklist

We used the 35 BMJ checklist items (hereafter: “items”) to assess included studies. Items were classified as not met (0), partially met (1), fully met (2), or not relevant (N/R). If relevant information was not contained in the primary publication or supplemental materials, but an appropriate citation was listed, we classified that item as “not available” (N/A). A detailed rubric was developed for each checklist item (**Appendix 4**). Average quality values were calculated for each question by summing the 0s, 1s, and 2s each article received across all studies, then dividing that sum by the number of items for which 0s, 1s, and 2s were possible (excluding N/R and N/A).

Additional items were created to track, in more detail, important article features which the BMJ checklist did not directly address but have been recommended in other authoritative guidelines (**Appendix 5**).^3, 7, 28^ These features did not contribute to average checklist values. During analysis, we grouped these additional items, along with select BMJ checklist items that we wished to highlight in more detail, into three distinct methodological constructs: perspective, scope, and parameter selection; the use of sensitivity/uncertainty analyses; and reporting transparency.

### Review Process

Article coding and assessment began with a “primary coder” who applied qualitative codes and assessed items. Next, a “secondary coder” received the already-coded articles from primary coders and cross-examined articles to ensure codes were appropriately applied. Secondary coders independently assessed all 35 BMJ items, and were blind to the assessment given by primary coders. Conflicts were discussed and resolved between the two reviewers (KJ, IG, KH, KHL).

## RESULTS

### Study Characteristics

Of the 5727 records identified through database searches, 47 studies met inclusion criteria (Figure 1). **Table 1** reports the main features of the 47 articles included in this review along with each article’s average quality assessment. Three genetic conditions constituted nearly half of all studies: Lynch syndrome (n=10), familial hypercholesterolemia (n = 7), and hereditary breast and ovarian cancer (n=14). A smaller set of studies considered maturity onset diabetes of the young (n=3), thrombophilia (n=2), or multiple conditions (n=2), and undiagnosed pediatric disorders (n=4). The setting of most studies was the United States (n=11), the United Kingdom (n=9), and Australia (n=9), with smaller numbers also conducted in Germany (n=4), both the United States and the United Kingdom (n=3), the Netherlands (n=3) and elsewhere (Spain: n=2; Poland: n=1; Norway: n=1; Malaysia: n=1; Italy: n=2; Taiwan, n=1; Singapore: n=1).

**Figure 1.**
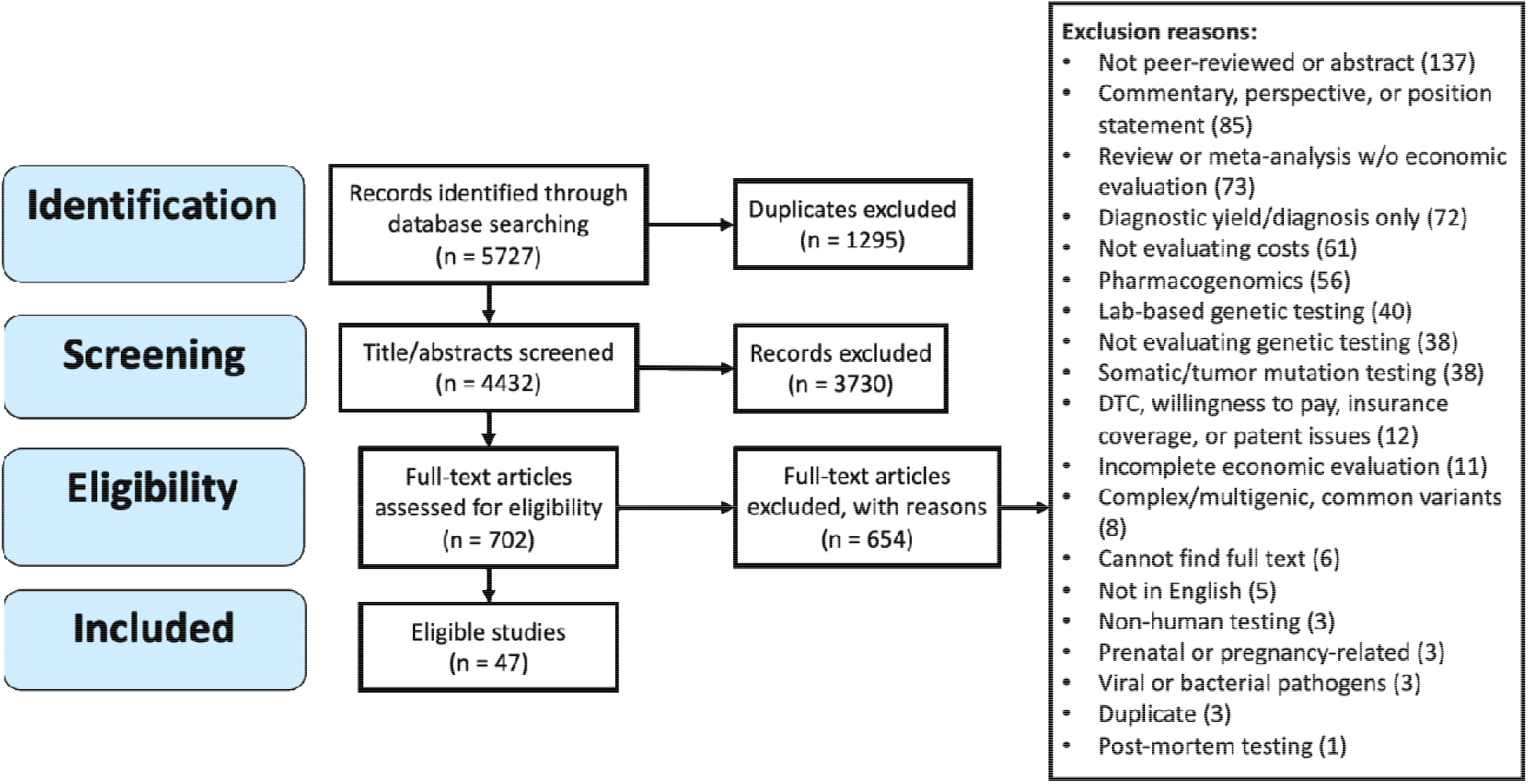
PRISMA search and exclusion flowchart.

**Table 1:**
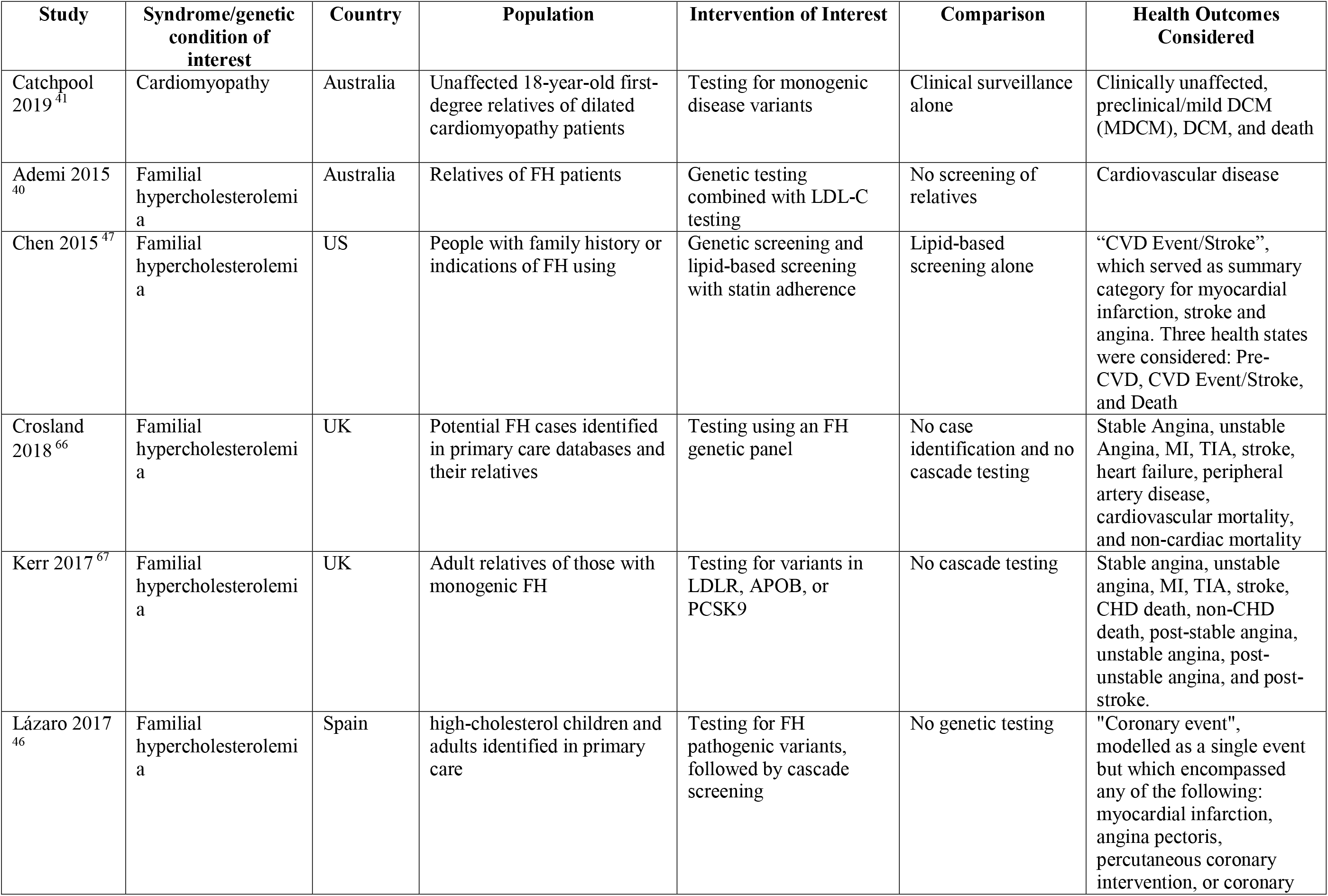
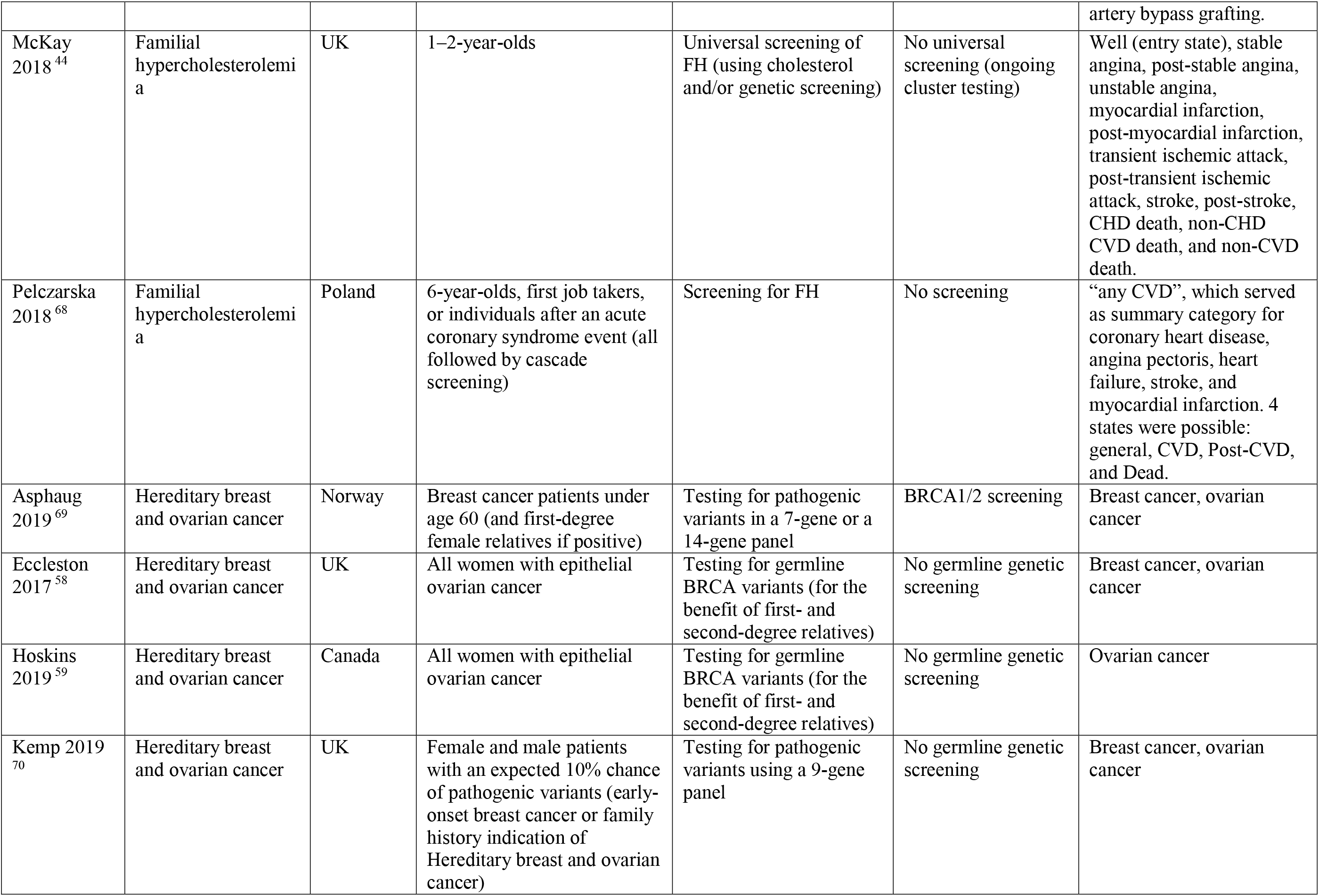
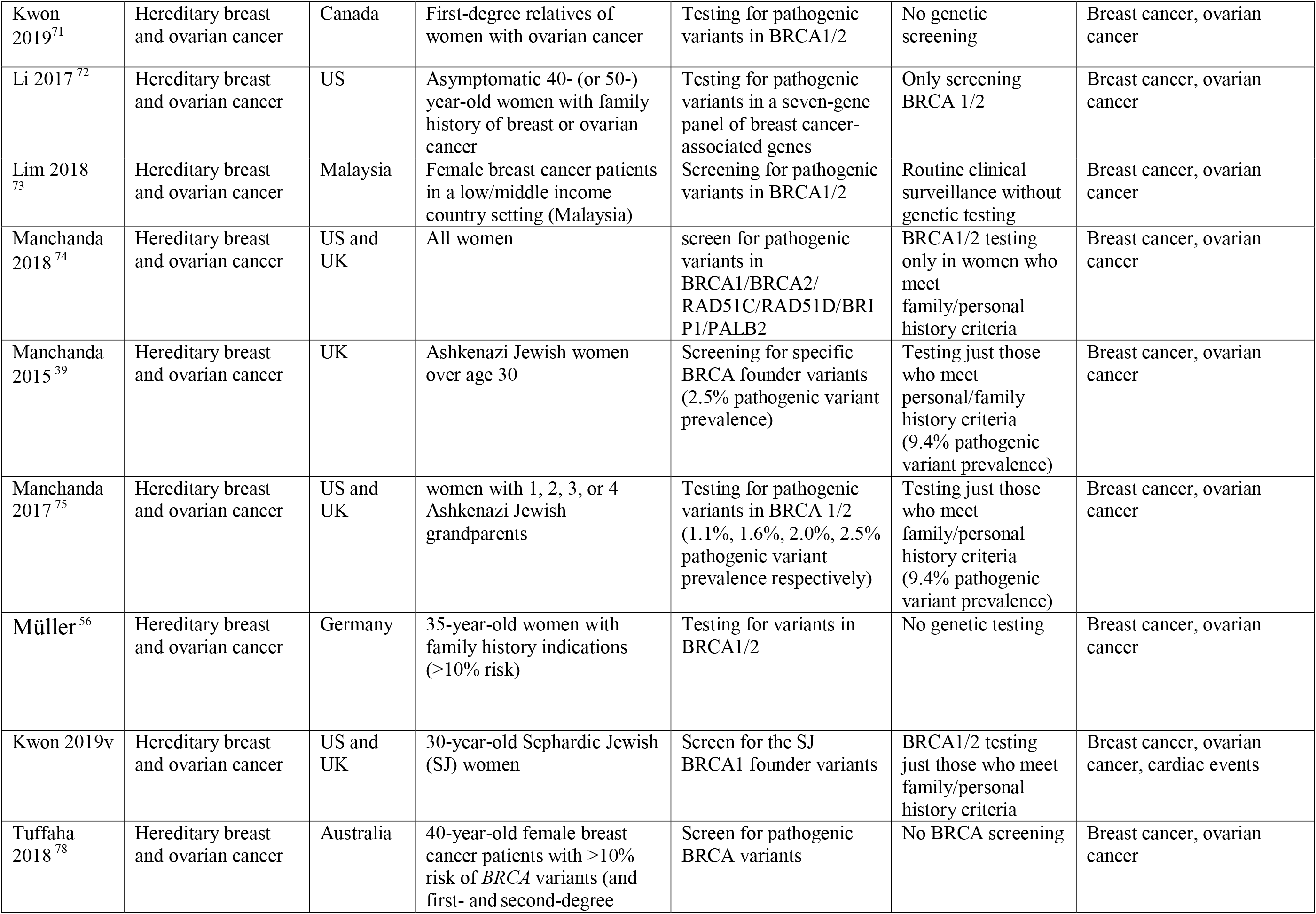
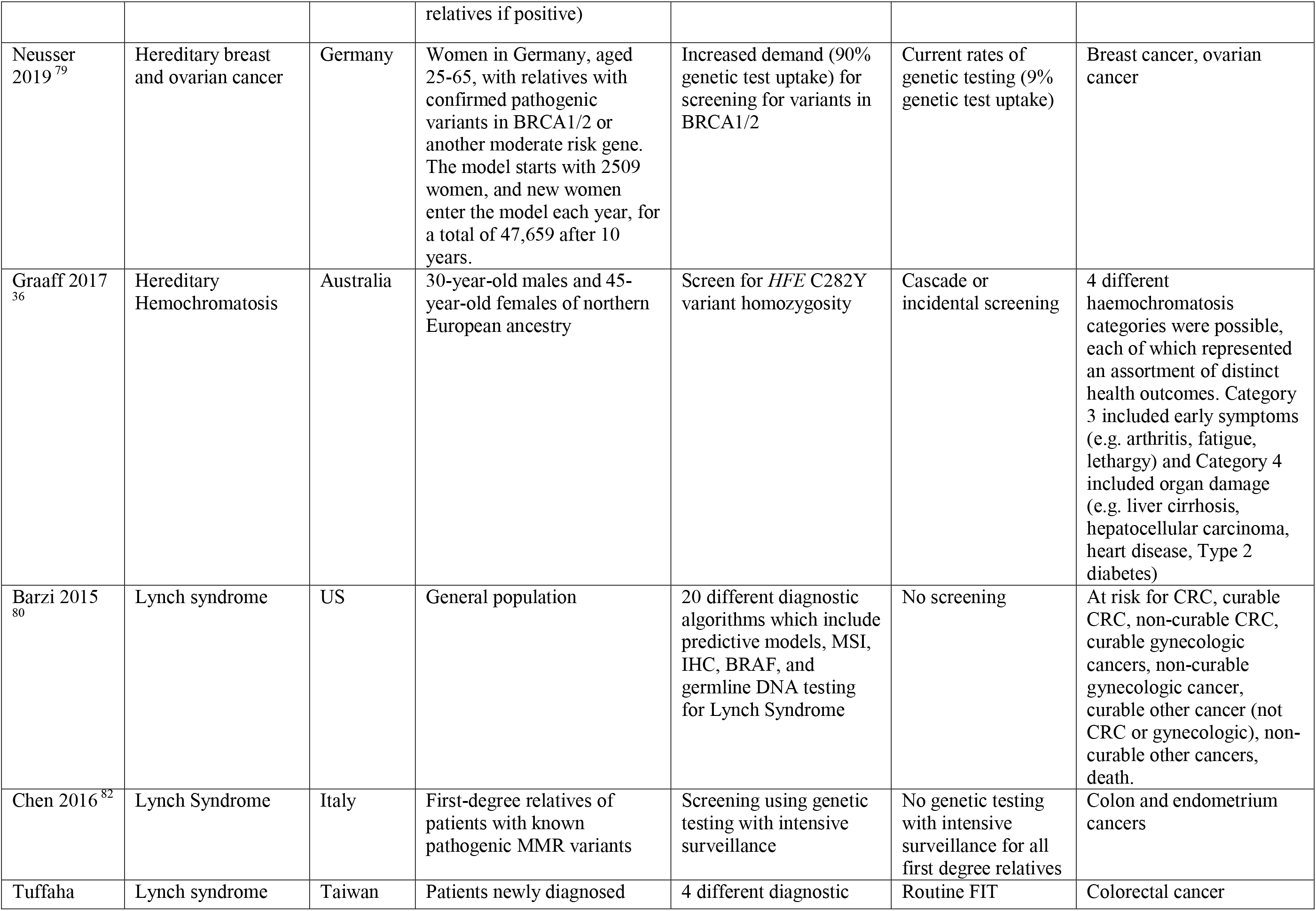
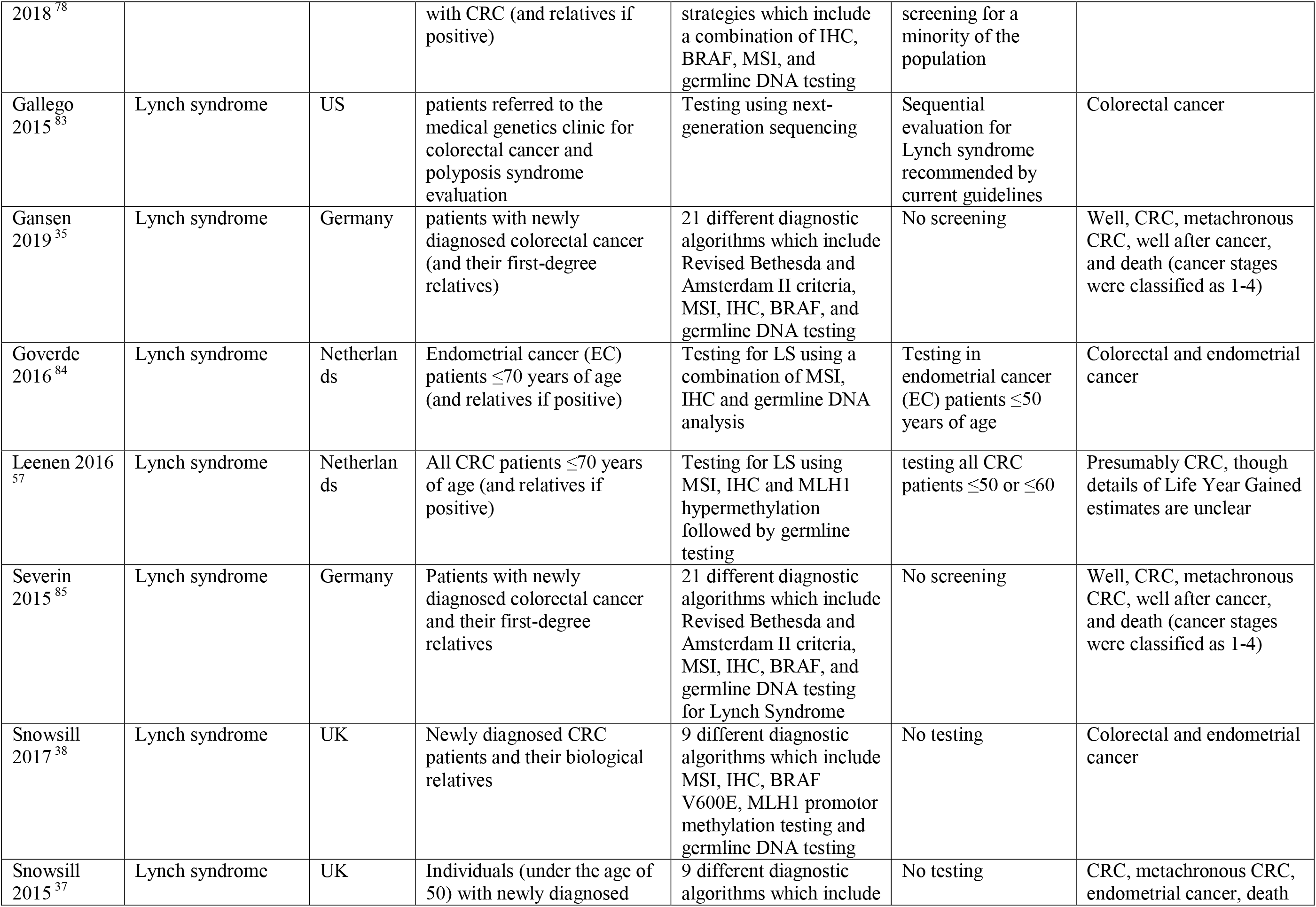
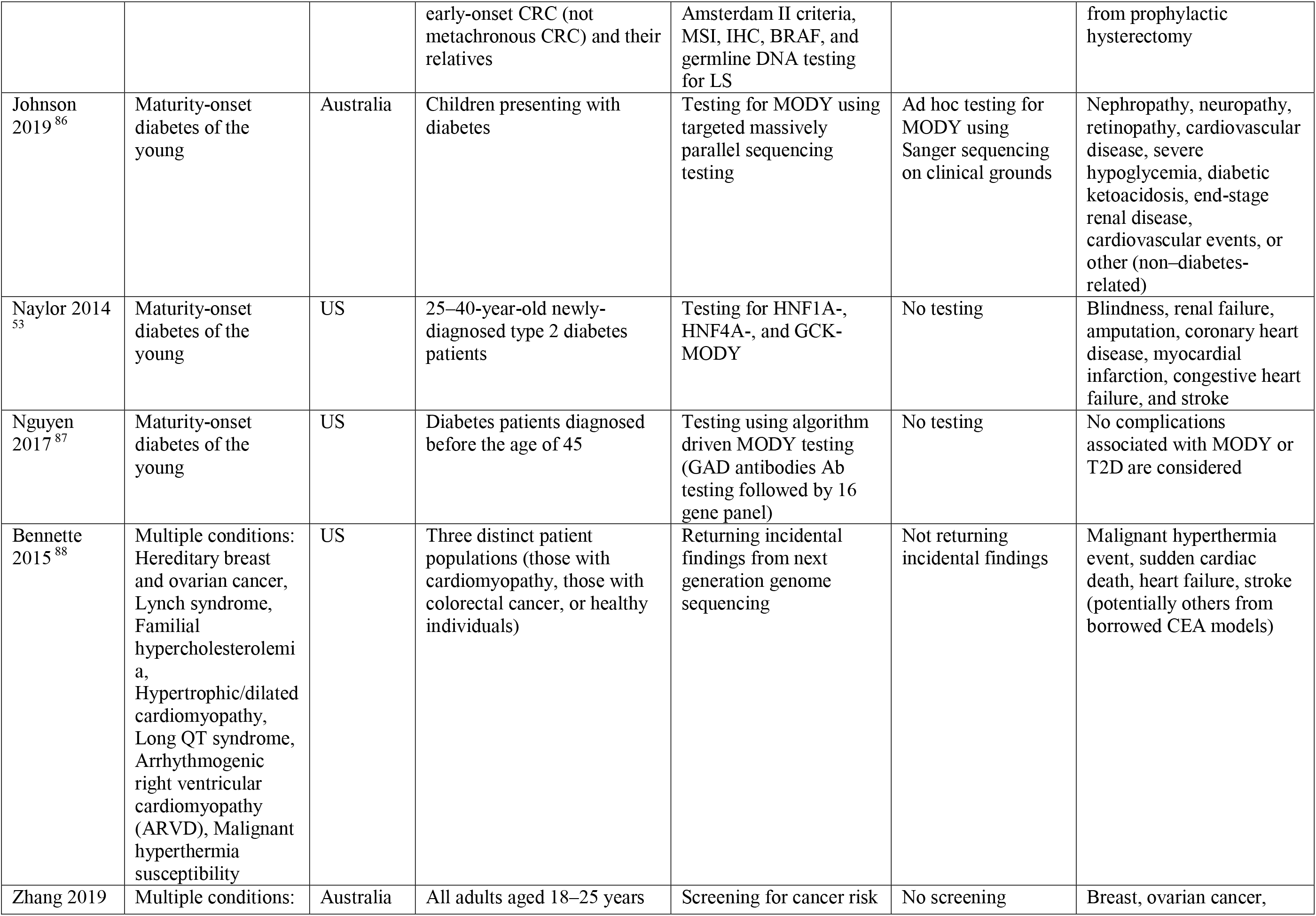
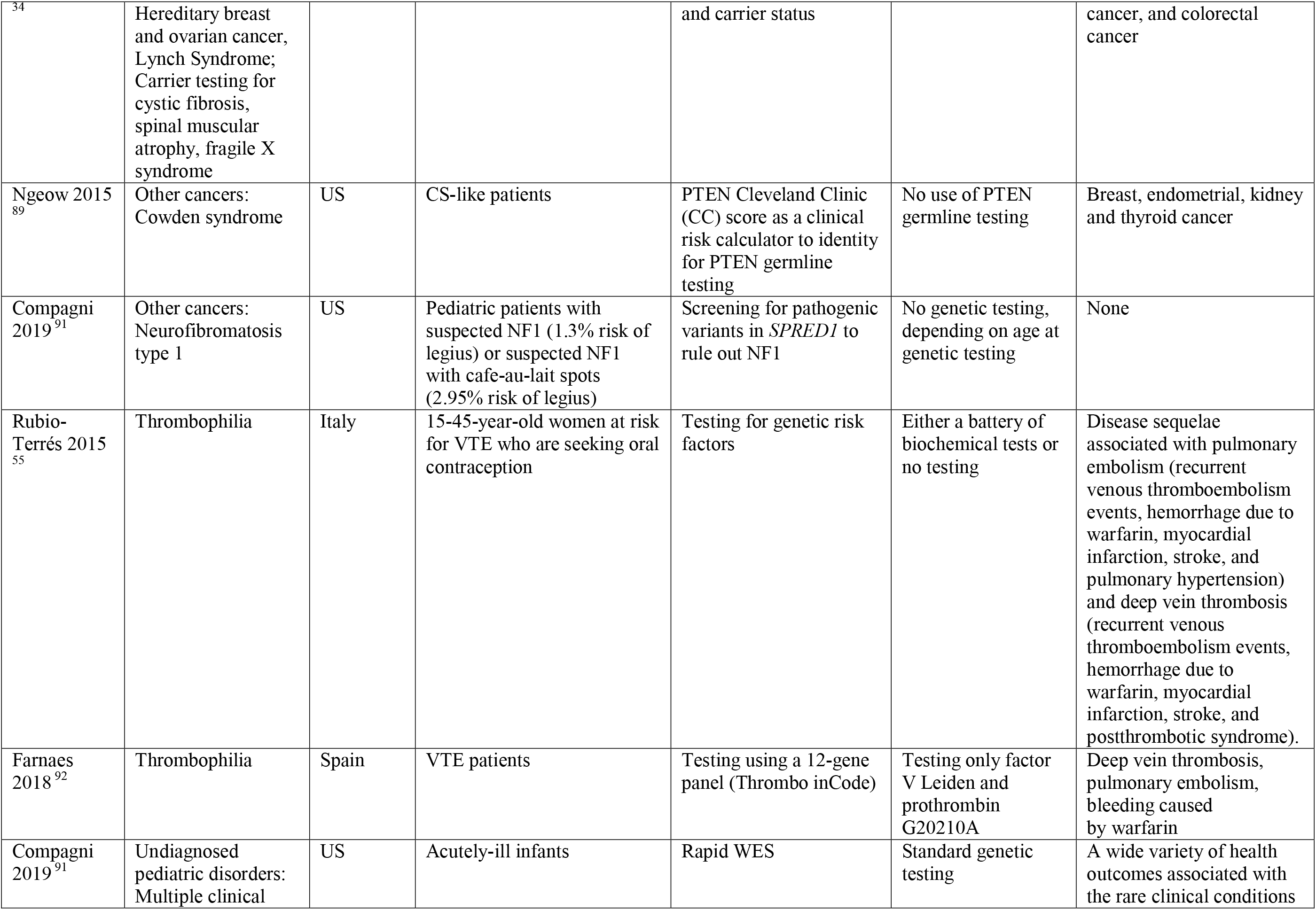
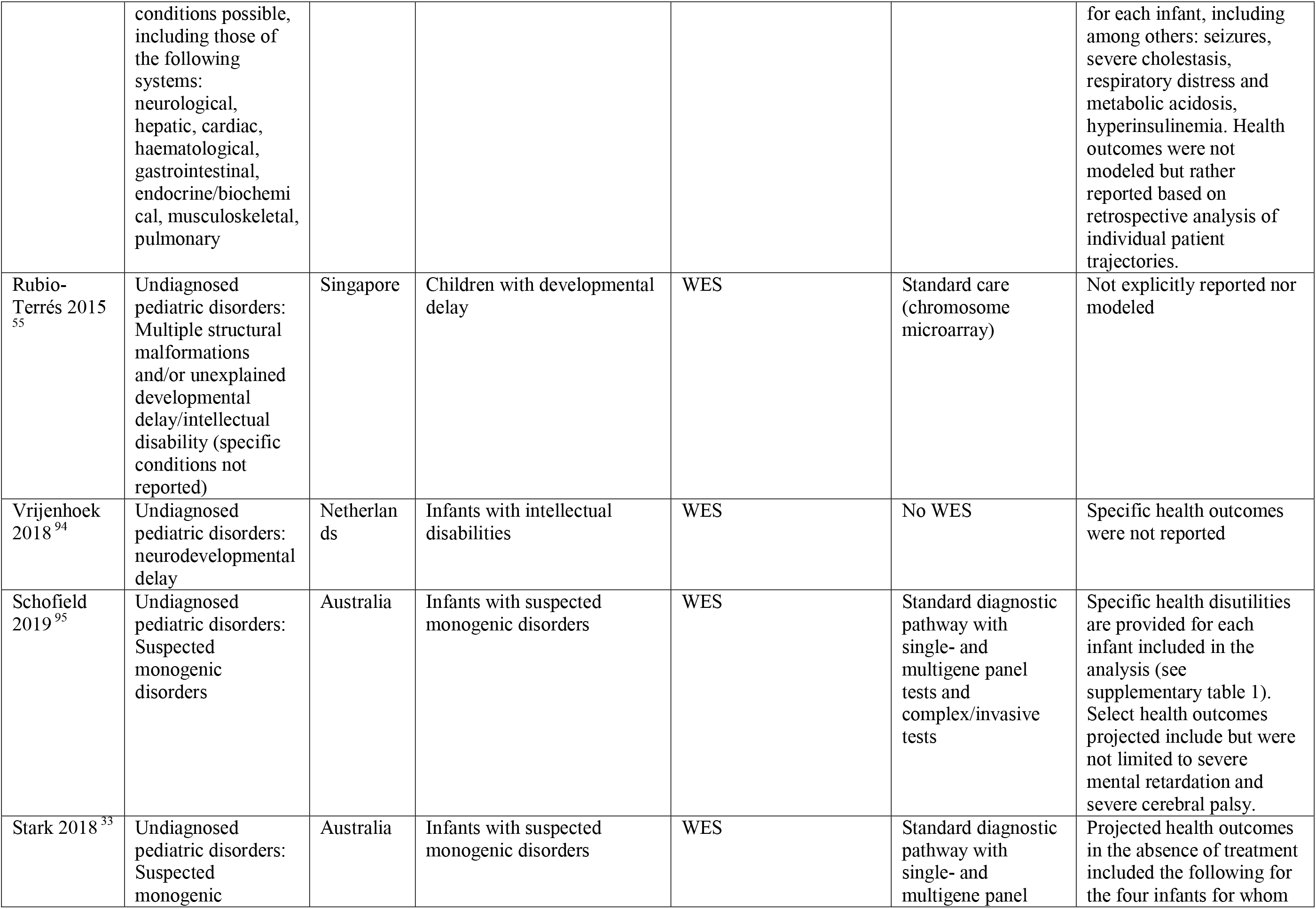
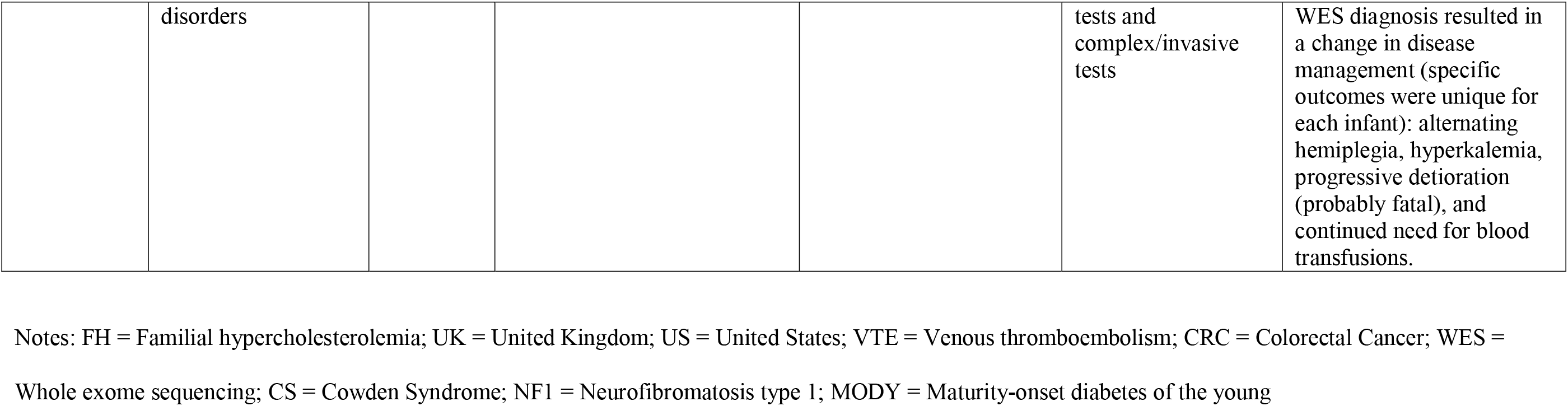
Summary Characteristics of Included Studies.

| Table 1: Summary Characteristics of Included Studies |  |  |  |  |  |  |
| --- | --- | --- | --- | --- | --- | --- |
| Study | Syndrome/genetic condition of interest | Country | Population | Intervention of Interest | Comparison | Health Outcomes Considered |
| Catchpool 2019<sup>41</sup> | Cardiomyopathy | Australia | Unaffected 18-year-old first-degree relatives of dilated cardiomyopathy patients | Testing for monogenic disease variants | Clinical surveillance alone | Clinically unaffected, preclinical/mild DCM (MDCM), DCM, and death |
| Ademi 2015<sup>40</sup> | Familial hypercholesterolemia | Australia | Relatives of FH patients | Genetic testing combined with LDL-C testing | No screening of relatives | Cardiovascular disease |
| Chen 2015<sup>47</sup> | Familial hypercholesterolemia | US | People with family history or indications of FH using | Genetic screening and lipid-based screening with statin adherence | Lipid-based screening alone | "CVD Event/Stroke", which served as summary category for myocardial infarction, stroke and angina. Three health states were considered: Pre-CVD, CVD Event/Stroke, and Death |
| Crosland 2018<sup>66</sup> | Familial hypercholesterolemia | UK | Potential FH cases identified in primary care databases and their relatives | Testing using an FH genetic panel | No case identification and no cascade testing | Stable Angina, unstable Angina, MI, TIA, stroke, heart failure, peripheral artery disease, cardiovascular mortality, and non-cardiac mortality |
| Kerr 2017<sup>67</sup> | Familial hypercholesterolemia | UK | Adult relatives of those with monogenic FH | Testing for variants in LDLR, APOB, or PCSK9 | No cascade testing | Stable angina, unstable angina, MI, TIA, stroke, CHD death, non-CHD death, post-stable angina, unstable angina, post-unstable angina, and post-stroke. |
| Lázaro 2017<sup>46</sup> | Familial hypercholesterolemia | Spain | high-cholesterol children and adults identified in primary care | Testing for FH pathogenic variants, followed by cascade screening | No genetic testing | "Coronary event", modelled as a single event but which encompassed any of the following: myocardial infarction, angina pectoris, percutaneous coronary intervention, or coronary artery bypass grafting. |
| McKay<br>2018<sup>44</sup> | Familial<br>hypercholesterolemia | UK | 1–2-year-olds | Universal screening of<br>FH (using cholesterol<br>and/or genetic screening) | No universal<br>screening (ongoing<br>cluster testing) | Well (entry state), stable<br>angina, post-stable angina,<br>unstable angina,<br>myocardial infarction,<br>post-myocardial infarction,<br>transient ischemic attack,<br>post-transient ischemic<br>attack, stroke, post-stroke,<br>CHD death, non-CHD<br>CVD death, and non-CVD<br>death. |
| Pelczarska<br>2018<sup>68</sup> | Familial<br>hypercholesterolemia | Poland | 6-year-olds, first job takers,<br>or individuals after an acute<br>coronary syndrome event (all<br>followed by cascade<br>screening) | Screening for FH | No screening | “any CVD”, which served<br>as summary category for<br>coronary heart disease,<br>angina pectoris, heart<br>failure, stroke, and<br>myocardial infarction. 4<br>states were possible:<br>general, CVD, Post-CVD,<br>and Dead. |
| Asphaug<br>2019<sup>69</sup> | Hereditary breast<br>and ovarian cancer | Norway | Breast cancer patients under<br>age 60 (and first-degree<br>female relatives if positive) | Testing for pathogenic<br>variants in a 7-gene or a<br>14-gene panel | BRCA1/2 screening | Breast cancer, ovarian<br>cancer |
| Eccleston<br>2017<sup>58</sup> | Hereditary breast<br>and ovarian cancer | UK | All women with epithelial<br>ovarian cancer | Testing for germline<br>BRCA variants (for the<br>benefit of first- and<br>second-degree relatives) | No germline genetic<br>screening | Breast cancer, ovarian<br>cancer |
| Hoskins<br>2019<sup>59</sup> | Hereditary breast<br>and ovarian cancer | Canada | All women with epithelial<br>ovarian cancer | Testing for germline<br>BRCA variants (for the<br>benefit of first- and<br>second-degree relatives) | No germline genetic<br>screening | Ovarian cancer |
| Kemp 2019<sup>70</sup> | Hereditary breast<br>and ovarian cancer | UK | Female and male patients<br>with an expected 10% chance<br>of pathogenic variants (early-onset breast cancer or family<br>history indication of<br>Hereditary breast and ovarian<br>cancer) | Testing for pathogenic<br>variants using a 9-gene<br>panel | No germline genetic<br>screening | Breast cancer, ovarian<br>cancer |
| Kwon 2019<sup>71</sup> | Hereditary breast and ovarian cancer | Canada | First-degree relatives of women with ovarian cancer | Testing for pathogenic variants in BRCA1/2 | No genetic screening | Breast cancer, ovarian cancer |
| Li 2017<sup>72</sup> | Hereditary breast and ovarian cancer | US | Asymptomatic 40- (or 50-) year-old women with family history of breast or ovarian cancer | Testing for pathogenic variants in a seven-gene panel of breast cancer-associated genes | Only screening BRCA 1/2 | Breast cancer, ovarian cancer |
| Lim 2018<sup>73</sup> | Hereditary breast and ovarian cancer | Malaysia | Female breast cancer patients in a low/middle income country setting (Malaysia) | Screening for pathogenic variants in BRCA1/2 | Routine clinical surveillance without genetic testing | Breast cancer, ovarian cancer |
| Manchanda 2018<sup>74</sup> | Hereditary breast and ovarian cancer | US and UK | All women | screen for pathogenic variants in BRCA1/BRCA2/RAD51C/RAD51D/BRI P1/PALB2 | BRCA1/2 testing only in women who meet family/personal history criteria | Breast cancer, ovarian cancer |
| Manchanda 2015<sup>39</sup> | Hereditary breast and ovarian cancer | UK | Ashkenazi Jewish women over age 30 | Screening for specific BRCA founder variants (2.5% pathogenic variant prevalence) | Testing just those who meet personal/family history criteria (9.4% pathogenic variant prevalence) | Breast cancer, ovarian cancer |
| Manchanda 2017<sup>75</sup> | Hereditary breast and ovarian cancer | US and UK | women with 1, 2, 3, or 4 Ashkenazi Jewish grandparents | Testing for pathogenic variants in BRCA 1/2 (1.1%, 1.6%, 2.0%, 2.5% pathogenic variant prevalence respectively) | Testing just those who meet family/personal history criteria (9.4% pathogenic variant prevalence) | Breast cancer, ovarian cancer |
| Müller<sup>56</sup> | Hereditary breast and ovarian cancer | Germany | 35-year-old women with family history indications (>10% risk) | Testing for variants in BRCA1/2 | No genetic testing | Breast cancer, ovarian cancer |
| Kwon 2019v | Hereditary breast and ovarian cancer | US and UK | 30-year-old Sephardic Jewish (SJ) women | Screen for the SJ BRCA1 founder variants | BRCA1/2 testing just those who meet family/personal history criteria | Breast cancer, ovarian cancer, cardiac events |
| Tuffaha 2018<sup>78</sup> | Hereditary breast and ovarian cancer | Australia | 40-year-old female breast cancer patients with >10% risk of *BRCA* variants (and first- and second-degree relatives if positive) | Screen for pathogenic BRCA variants | No BRCA screening | Breast cancer, ovarian cancer |
| Neusser 2019<sup>79</sup> | Hereditary breast and ovarian cancer | Germany | Women in Germany, aged 25-65, with relatives with confirmed pathogenic variants in BRCA1/2 or another moderate risk gene. The model starts with 2509 women, and new women enter the model each year, for a total of 47,659 after 10 years. | Increased demand (90% genetic test uptake) for screening for variants in BRCA1/2 | Current rates of genetic testing (9% genetic test uptake) | Breast cancer, ovarian cancer |
| Graaff 2017<sup>36</sup> | Hereditary Hemochromatosis | Australia | 30-year-old males and 45-year-old females of northern European ancestry | Screen for *HFE* C282Y variant homozygosity | Cascade or incidental screening | 4 different haemochromatosis categories were possible, each of which represented an assortment of distinct health outcomes. Category 3 included early symptoms (e.g. arthritis, fatigue, lethargy) and Category 4 included organ damage (e.g. liver cirrhosis, hepatocellular carcinoma, heart disease, Type 2 diabetes) |
| Barzi 2015<sup>80</sup> | Lynch syndrome | US | General population | 20 different diagnostic algorithms which include predictive models, MSI, IHC, BRAF, and germline DNA testing for Lynch Syndrome | No screening | At risk for CRC, curable CRC, non-curable CRC, curable gynecologic cancers, non-curable gynecologic cancer, curable other cancer (not CRC or gynecologic), non-curable other cancers, death. |
| Chen 2016<sup>82</sup> | Lynch Syndrome | Italy | First-degree relatives of patients with known pathogenic MMR variants | Screening using genetic testing with intensive surveillance | No genetic testing with intensive surveillance for all first degree relatives | Colon and endometrium cancers |
| Tuffaha | Lynch syndrome | Taiwan | Patients newly diagnosed | 4 different diagnostic | Routine FIT | Colorectal cancer |
| 2018<sup>78</sup> |  |  | with CRC (and relatives if positive) | strategies which include a combination of IHC, BRAF, MSI, and germline DNA testing | screening for a minority of the population |  |
| Gallego 2015<sup>83</sup> | Lynch syndrome | US | patients referred to the medical genetics clinic for colorectal cancer and polyposis syndrome evaluation | Testing using next-generation sequencing | Sequential evaluation for Lynch syndrome recommended by current guidelines | Colorectal cancer |
| Gansen 2019<sup>35</sup> | Lynch syndrome | Germany | patients with newly diagnosed colorectal cancer (and their first-degree relatives) | 21 different diagnostic algorithms which include Revised Bethesda and Amsterdam II criteria, MSI, IHC, BRAF, and germline DNA testing | No screening | Well, CRC, metachronous CRC, well after cancer, and death (cancer stages were classified as 1-4) |
| Goverde 2016<sup>84</sup> | Lynch syndrome | Netherlands | Endometrial cancer (EC) patients $\leq 70$ years of age (and relatives if positive) | Testing for LS using a combination of MSI, IHC and germline DNA analysis | Testing in endometrial cancer (EC) patients $\leq 50$ years of age | Colorectal and endometrial cancer |
| Leenen 2016<sup>57</sup> | Lynch syndrome | Netherlands | All CRC patients $\leq 70$ years of age (and relatives if positive) | Testing for LS using MSI, IHC and MLH1 hypermethylation followed by germline testing | testing all CRC patients $\leq 50$ or $\leq 60$ | Presumably CRC, though details of Life Year Gained estimates are unclear |
| Severin 2015<sup>85</sup> | Lynch syndrome | Germany | Patients with newly diagnosed colorectal cancer and their first-degree relatives | 21 different diagnostic algorithms which include Revised Bethesda and Amsterdam II criteria, MSI, IHC, BRAF, and germline DNA testing for Lynch Syndrome | No screening | Well, CRC, metachronous CRC, well after cancer, and death (cancer stages were classified as 1-4) |
| Snowsill 2017<sup>38</sup> | Lynch syndrome | UK | Newly diagnosed CRC patients and their biological relatives | 9 different diagnostic algorithms which include MSI, IHC, BRAF V600E, MLH1 promotor methylation testing and germline DNA testing | No testing | Colorectal and endometrial cancer |
| Snowsill 2015<sup>37</sup> | Lynch syndrome | UK | Individuals (under the age of 50) with newly diagnosed | 9 different diagnostic algorithms which include | No testing | CRC, metachronous CRC, endometrial cancer, death |
|  |  |  | early-onset CRC (not metachronous CRC) and their relatives | Amsterdam II criteria, MSI, IHC, BRAF, and germline DNA testing for LS |  | from prophylactic hysterectomy |
| Johnson 2019<sup>86</sup> | Maturity-onset diabetes of the young | Australia | Children presenting with diabetes | Testing for MODY using targeted massively parallel sequencing testing | Ad hoc testing for MODY using Sanger sequencing on clinical grounds | Nephropathy, neuropathy, retinopathy, cardiovascular disease, severe hypoglycemia, diabetic ketoacidosis, end-stage renal disease, cardiovascular events, or other (non–diabetes-related) |
| Naylor 2014<sup>53</sup> | Maturity-onset diabetes of the young | US | 25–40-year-old newly-diagnosed type 2 diabetes patients | Testing for HNF1A-, HNF4A-, and GCK-MODY | No testing | Blindness, renal failure, amputation, coronary heart disease, myocardial infarction, congestive heart failure, and stroke |
| Nguyen 2017<sup>87</sup> | Maturity-onset diabetes of the young | US | Diabetes patients diagnosed before the age of 45 | Testing using algorithm driven MODY testing (GAD antibodies Ab testing followed by 16 gene panel) | No testing | No complications associated with MODY or T2D are considered |
| Bennette 2015<sup>88</sup> | Multiple conditions: Hereditary breast and ovarian cancer, Lynch syndrome, Familial hypercholesterolemia, Hypertrophic/dilated cardiomyopathy, Long QT syndrome, Arrhythmogenic right ventricular cardiomyopathy (ARVD), Malignant hyperthermia susceptibility | US | Three distinct patient populations (those with cardiomyopathy, those with colorectal cancer, or healthy individuals) | Returning incidental findings from next generation genome sequencing | Not returning incidental findings | Malignant hyperthermia event, sudden cardiac death, heart failure, stroke (potentially others from borrowed CEA models) |
| Zhang 2019 | Multiple conditions: | Australia | All adults aged 18–25 years | Screening for cancer risk | No screening | Breast, ovarian cancer, |
| <sup>34</sup> | Hereditary breast and ovarian cancer, Lynch Syndrome; Carrier testing for cystic fibrosis, spinal muscular atrophy, fragile X syndrome |  |  | and carrier status |  | cancer, and colorectal cancer |
| Ngeow 2015<sup>89</sup> | Other cancers: Cowden syndrome | US | CS-like patients | PTEN Cleveland Clinic (CC) score as a clinical risk calculator to identify for PTEN germline testing | No use of PTEN germline testing | Breast, endometrial, kidney and thyroid cancer |
| Compagni 2019<sup>91</sup> | Other cancers: Neurofibromatosis type 1 | US | Pediatric patients with suspected NF1 (1.3% risk of legius) or suspected NF1 with cafe-au-lait spots (2.95% risk of legius) | Screening for pathogenic variants in *SPRED1* to rule out NF1 | No genetic testing, depending on age at genetic testing | None |
| Rubio-Terrés 2015<sup>55</sup> | Thrombophilia | Italy | 15-45-year-old women at risk for VTE who are seeking oral contraception | Testing for genetic risk factors | Either a battery of biochemical tests or no testing | Disease sequelae associated with pulmonary embolism (recurrent venous thromboembolism events, hemorrhage due to warfarin, myocardial infarction, stroke, and pulmonary hypertension) and deep vein thrombosis (recurrent venous thromboembolism events, hemorrhage due to warfarin, myocardial infarction, stroke, and postthrombotic syndrome). |
| Farnaes 2018<sup>92</sup> | Thrombophilia | Spain | VTE patients | Testing using a 12-gene panel (Thrombo inCode) | Testing only factor V Leiden and prothrombin G20210A | Deep vein thrombosis, pulmonary embolism, bleeding caused by warfarin |
| Compagni 2019<sup>91</sup> | Undiagnosed pediatric disorders: Multiple clinical | US | Acutely-ill infants | Rapid WES | Standard genetic testing | A wide variety of health outcomes associated with the rare clinical conditions |
|  | conditions possible, including those of the following systems: neurological, hepatic, cardiac, haematological, gastrointestinal, endocrine/biochemical, musculoskeletal, pulmonary |  |  |  |  | for each infant, including among others: seizures, severe cholestasis, respiratory distress and metabolic acidosis, hyperinsulinemia. Health outcomes were not modeled but rather reported based on retrospective analysis of individual patient trajectories. |
| Rubio-Terrés 2015<sup>55</sup> | Undiagnosed pediatric disorders: Multiple structural malformations and/or unexplained developmental delay/intellectual disability (specific conditions not reported) | Singapore | Children with developmental delay | WES | Standard care (chromosome microarray) | Not explicitly reported nor modeled |
| Vrijenhoek 2018<sup>94</sup> | Undiagnosed pediatric disorders: neurodevelopmental delay | Netherlands | Infants with intellectual disabilities | WES | No WES | Specific health outcomes were not reported |
| Schofield 2019<sup>95</sup> | Undiagnosed pediatric disorders: Suspected monogenic disorders | Australia | Infants with suspected monogenic disorders | WES | Standard diagnostic pathway with single- and multigene panel tests and complex/invasive tests | Specific health disutilities are provided for each infant included in the analysis (see supplementary table 1). Select health outcomes projected include but were not limited to severe mental retardation and severe cerebral palsy. |
| Stark 2018<sup>33</sup> | Undiagnosed pediatric disorders: Suspected monogenic | Australia | Infants with suspected monogenic disorders | WES | Standard diagnostic pathway with single- and multigene panel | Projected health outcomes in the absence of treatment included the following for the four infants for whom |
|  | disorders |  |  |  | tests and complex/invasive tests | WES diagnosis resulted in a change in disease management (specific outcomes were unique for each infant): alternating hemiplegia, hyperkalemia, progressive deterioration (probably fatal), and continued need for blood transfusions. |

Notes: FH = Familial hypercholesterolemia; UK = United Kingdom; US = United States; VTE = Venous thromboembolism; CRC = Colorectal Cancer; WES = Whole exome sequencing; CS = Cowden Syndrome; NF1 = Neurofibromatosis type 1; MODY = Maturity-onset diabetes of the young

**Table 2** presents the major model characteristics across all studies. Most studies utilized the combination of a decision tree with a Markov model (n = 17), though several used either just a decision tree (n = 11) or just a Markov model (n = 6). Five studies employed some form of individual-level simulation (e.g., microsimulation). Less than half of all studies (n = 18) compared only one alternative to “usual care”, which often consisted of the standard-of-care genetic or clinical testing/screening protocol. Most studies conducted cost-utility analyses (i.e., health outcomes are expressed in utility measures like QALYs or DALYs) (CUAs) (n = 26), with several conducting both CUAs and cost-effectiveness analyses (i.e., health outcomes are expressed in clinical measures like total diagnoses or deaths) (CEAs) (n = 10) and a limited number (n=6) conducting cost-consequence analyses. Three studies incorporated societal costs, the rest were strictly from either the healthcare sector or payer perspective.

**Table 2:** Primary Modeling Characteristics of Included Studies.

| **Table 2: Primary Modeling Characteristics of Included Studies** |  |  |  |  |  |  |  |  |
| --- | --- | --- | --- | --- | --- | --- | --- | --- |
| **Study** | **Type of evaluation** | **Perspective** | **Discounting** | **Time Horizon** | **Model type** | **Costing method** | **Sensitivity Analyses Conducted** | **Forms of Analysis Presentation** |
| Catchpool 2019<sup>41</sup> | CUA | Healthcare system (Australian Government) | 5% costs and outcomes | Lifetime | Decision tree and Markov model | Gross | PSA, One/Two-Way Deterministic Analysis | CEAC, Tornado Diagram |
| Ademi 2015<sup>40</sup> | CEA and CUA | Healthcare system (Australian Government) | 5% costs and outcomes | 10 years | Decision tree and Markov model | Gross | PSA, One/Two-Way Deterministic Analysis | CE Plane/Scatter Plot, CEAC |
| Chen 2015<sup>47</sup> | CUA | Societal and healthcare system combined | 3% costs and outcomes | Lifetime | Decision tree and Markov model | Gross | Threshold Analysis, PSA, One/Two-Way Deterministic Analysis | CEAC, Tornado Diagram |
| Crosland 2018<sup>66</sup> | CUA | Healthcare system (UK NHS) | 3.5% costs and outcomes | Lifetime | Decision tree and Markov model | Micro | Threshold Analysis, PSA, One/Two-Way Deterministic Analysis | CE Plane/Scatter Plot, CEAC |
| Kerr 2017<sup>67</sup> | CUA | Healthcare system (UK NHS) | 3.5% costs and outcomes | 30 years | Markov model | Micro | One/Two-Way Deterministic Analysis | None |
| Lázaro 2017<sup>46</sup> | CEA and CUA | Healthcare system (Spanish National Health System) and | 3% costs and outcomes | 10 years | Decision tree | Gross | Scenario Analysis, One/Two-Way Deterministic Analysis | CE Frontier |
|  |  | Societal |  |  |  |  |  |  |
| McKay 2018<sup>44</sup> | CUA | Healthcare system (UK NHS) | 3.5% costs and outcomes | Lifetime (limited to 100 years) | Decision tree and Markov model | Micro | Threshold Analysis, PSA, One/Two-Way Deterministic Analysis | CEAC |
| Pelczarska 2018<sup>68</sup> | CEA and CUA | Healthcare system (Polish Government) | 5% costs, 3.5% outcomes | Lifetime | Decision tree and Markov model | Gross | One/Two-Way Deterministic Analysis | None |
| Asphaug 2019<sup>69</sup> | CUA | Healthcare sector | 4% costs and outcomes | Lifetime (limited to 100 years) | Patient-level microsimulation with memory. | Micro | PSA | CEAC |
| Eccleston 2017<sup>58</sup> | CUA | Healthcare system (UK NHS) | 3.5% costs and outcomes | 50 years | Patient-level microsimulation with memory | Gross | Scenario Analysis, PSA, One/Two-Way Deterministic Analysis | CE Plane/Scatter Plot, CEAC |
| Hoskins 2019<sup>59</sup> | CUA | Canadian healthcare system perspective | 1.5% costs and outcomes | 50 years | Patient-level microsimulation with memory | Gross | Scenario Analysis, PSA, One/Two-Way Deterministic Analysis | None |
| Kemp 2019<sup>70</sup> | CUA | Healthcare system (UK NHS) | 3.5% costs and outcomes | 50 years | Patient-level microsimulation with memory | Gross | PSA | CEAC |
| Kwon 2019 | CUA | Healthcare system (Canadian Government) | 3% costs and outcomes | 50 years | Decision tree and Markov model | Gross | Threshold Analysis, Scenario Analysis, One/Two-Way Deterministic Analysis | None |
| Li 2017<sup>72</sup> | CEA and CUA | Healthcare payer | 3.5% costs and outcomes (strictly for QALYs, not life-years) | Lifetime (limited to 100 years) | Decision tree and Markov model | Gross | PSA, One/Two-Way Deterministic Analysis | CE Plane/Scatter Plot, CEAC, Tornado Diagram |
| Lim 2018<sup>73</sup> | CEA and CUA | Healthcare system (Malaysian Government) | 3% costs and outcomes | Lifetime | Decision tree and Markov model | Gross | Scenario Analysis, PSA, One/Two-Way Deterministic Analysis | CEAC, Tornado Diagram |
| Manchanda 2018<sup>74</sup> | CEA and CUA | Healthcare system (US and UK) | 3.5% costs and outcomes | Lifetime (to age 83 based on life tables) | Decision tree | Gross | Scenario Analysis, PSA, One/Two-Way Deterministic Analysis | CEAC |
| Manchanda | CUA | Healthcare | 3.5% cost | Lifetime (to | Decision tree | Gross | Scenario Analysis, PSA, | CEAC, |
| 2015<sup>39</sup> |  | system (UK NHS) | and outcomes | age 83 based on life tables) |  |  | One/Two-Way Deterministic Analysis | Tornado Diagram |
| Manchanda 2017<sup>75</sup> | CEA and CUA | Healthcare system (US and UK) | 3.5% costs and outcomes | Lifetime (to age 83 based on life tables) | Decision tree | Gross | Scenario Analysis, PSA, | CEAC |
| Müller<sup>56</sup> | CEA and CUA | Healthcare payer (German Statutory Health Insurance) | 3% costs and outcomes | 65 years | Decision tree and Markov model | Gross | PSA, One/Two-Way Deterministic Analysis | CEAC |
| Patel 2018<sup>77</sup> | CEA and CUA | Healthcare payer | 3.5% costs and outcomes | Lifetime (up until 83 and 82 years for UK and US women, respectively) | Markov model | Gross | Scenario Analysis, PSA, One/Two-Way Deterministic Analysis | CEAC, Tornado Diagram |
| Tuffaha 2018<sup>78</sup> | CUA | Healthcare system (Australian Government) | 5% costs and outcomes | Lifetime (limited to 90 years) | Decision tree and Markov model | Gross | PSA, One/Two-Way Deterministic Analysis | None |
| Neusser 2019<sup>79</sup> | CCA | Healthcare payer (German Statutory Health Insurance) | 3% costs | 10 years | Markov model | Gross | None | None |
| Graaff 2017<sup>36</sup> | CUA | Healthcare system (Australian Government) | 5% costs and outcomes | Lifetime | Markov model | Micro | PSA, One/Two-Way Deterministic Analysis | CEAC |
| Barzi 2015<sup>80</sup> | CEA | Societal (no clear societal costs) and healthcare system | 3% costs and outcomes | Whichever comes first: the death, an age of 80 years, or 50 | Decision tree followed by Markov -based individual patient simulation | Micro | Scenario Analysis, One/Two-Way Deterministic Analysis | None |
|  |  | combined |  | years of follow-up. |  |  |  |  |
| Bonfanti 2016<sup>81</sup> | CCA | Not stated (assumed healthcare system) | **Discounted at the 2012 level | 10 years | Informal epidemiological model | Micro | None | None |
| Chen 2016<sup>82</sup> | CEA | Healthcare system (The Ministry of Health and Welfare (MOHW) of the Taiwan government) | 3% costs and outcomes | Lifetime | Decision tree and Markov model | Gross | PSA, One/Two-Way Deterministic Analysis | CEAC, Tornado Diagram |
| Gallego 2015<sup>83</sup> | CUA and CEA (exclusively CUA in sensitivity analyses) | Not stated (assumed healthcare payer) | 3% (unclear how applied) | Lifetime | Decision tree | Gross | Scenario Analysis, PSA, One/Two-Way Deterministic Analysis | CEAC, Tornado Diagram |
| Gansen 2019<sup>35</sup> | CEA | Healthcare payer (German Statutory Health Insurance) | 3% costs and outcomes | 120 years | Decision tree and Markov model | Micro | Scenario Analysis, PSA | CE Frontier |
| Goverde 2016<sup>84</sup> | CEA | Not stated | 3% costs and outcomes | Not stated (presumably lifetime) | Decision tree | Micro | One/Two-Way Deterministic Analysis | Tornado Diagram |
| Leenen 2016<sup>57</sup> | CEA | Healthcare sector | 3% costs and outcomes | Lifetime | Decision tree | Micro | None | Tornado Diagram |
| Severin 2015<sup>85</sup> | CEA | Healthcare payer (German Statutory Health | 3% costs and outcomes | Lifetime (limited to 120 years) | Decision tree and Markov model | Micro | Scenario Analysis, PSA, One/Two-Way Deterministic Analysis | CEAC, Tornado Diagram |
|  |  | Insurance) |  |  |  |  |  |  |
| Snowsill 2017<sup>38</sup> | CUA | Healthcare system (UK NHS) and Personal Social Service | 3.5% costs and outcomes (strictly for QALYs, not life-years) | Lifetime (limited to 100 years) | Decision tree and individual patient simulation | Micro | Scenario Analysis | CE Frontier |
| Snowsill 2015<sup>37</sup> | CUA | Healthcare system (UK NHS) | 3.5% costs and outcomes | Lifetime (limited to 100 years) | Decision tree and individual patient simulation | Micro | Scenario Analysis, One/Two-Way Deterministic Analysis | CE Frontier, Tornado Diagram |
| Johnson 2019<sup>86</sup> | CUA | Healthcare system (Australian Government) | 3% costs and outcomes | 10 years and 30 years | Decision tree and Markov model | Gross | One/Two-Way Deterministic Analysis | Tornado Diagram |
| Naylor 2014<sup>53</sup> | CUA | Healthcare system | 3% costs and outcomes | Lifetime | Decision tree followed by Markov-based individual patient simulation | Gross | Threshold Analysis, Scenario Analysis, One/Two-Way Deterministic Analysis | Tornado Diagram |
| Nguyen 2017<sup>87</sup> | CUA | Healthcare payer | 3.5% costs and outcomes | 30 years | Decision tree | Gross | Threshold Analysis, PSA, One/Two-Way Deterministic Analysis | CEAC, Tornado Diagram |
| Bennette 2015<sup>88</sup> | CUA | Healthcare system | 3% costs and outcomes | Lifetime | Decision tree and Markov model | Gross (costs based on prior CEAs) | Threshold Analysis, Scenario Analysis, PSA, One/Two-Way Deterministic Analysis | CEAC |
| Zhang 2019<sup>34</sup> | CUA | Healthcare system | 3% costs and outcomes | Lifetime | Decision tree | Gross | Scenario Analysis, PSA, One/Two-Way Deterministic Analysis | CE Plane/Scatter Plot |
| Ngeow 2015<sup>89</sup> | CUA | Societal and healthcare system combined | 3% costs and outcomes | Lifetime | Decision tree and Markov model | Gross | Scenario Analysis, PSA, | CEAC, Tornado Diagram |
| Muram 2013<sup>90</sup> | CCA | Healthcare payer | 3% costs and outcomes | 17 years (18 months old - 18 years old) | Markov model and individual patient simulation | Gross | None | None |
| Compagni 2019<sup>91</sup> | CUA | Healthcare system (Italian National Health System) | 3.5% costs and benefits | Lifetime | Decision tree | Micro | Scenario Analysis, PSA, One/Two-Way Deterministic Analysis | CE Plane/Scatter Plot, CEAC, Tornado Diagram |
| Rubio-Terrés 2015<sup>55</sup> | CUA | Healthcare system (UK NHS) | 3.5% costs and outcomes | 35 years | Decision tree | Gross | Threshold Analysis, Scenario Analysis, PSA, One/Two-Way Deterministic Analysis | CE Plane/Scatter Plot, Tornado Diagram |
| Farnaes 2018<sup>92</sup> | CCA | Healthcare system | N/A | Various for different infants | N/A | Gross | None | None |
| Hayeems 2017<sup>93</sup> | CCA | Healthcare system | N/A | On average, 15 months after diagnostic results (standard care or WGS) were reported. | Linear mixed effects model | Gross | None | None |
| Vrijenhoek 2018<sup>94</sup> | CCA | Healthcare system | N/A | The length of follow-up was, on average, 240 days after WES and 922 days before WES. | None | Micro | None | None |
| Schofield 2019 | CUA | Not stated | 5% (unclear how) | 20 years | Decision tree | Gross | One/Two-Way Deterministic Analysis | None |
| <sup>95</sup> |  |  | applied) |  |  |  |  |  |
| Stark 2018<sup>33</sup> | CUA | Healthcare system | None stated | Mediation duration of follow-up: 473 days (interquartile range: 411–650) | No formal model (individual prospective cohort) | Gross | PSA | CE Plane/Scatter Plot |

Notes: DMC = Dilated cardiomyopathy; CEA = Cost-Effectiveness Analysis; CE Plane = Cost-Effectiveness Plane; CEAC = Cost-Effectiveness Acceptability Curve; CE = Cost-Effectiveness Frontier; CUA = Cost-Utility Analysis; CCA = Const-Consequence Analysis; NHS = National Health Service; PSA = Probabilistic Sensitivity Analysis; UK = United Kingdom; US = United States

### BMJ Checklist Assessment

Some basic items from the BMJ checklist were fully met by nearly all studies, including “The research question is stated” (average value [AV]: 2) and “The primary outcome measure(s) for the economic evaluation are clearly stated” (AV: 1.98). Conversely, several checklist items consistently were not met or partially met by all studies, including “Quantities of resources are reported separately from their unit costs” (AV: 0.87) and “Details of the subjects from whom valuations were obtained are given” (AV: 1.10). Some items were consistently addressed by citing external sources but without an overview of the source material (N/A), such as “Methods to value health states and other benefits are stated.” Several of the cost-consequence analyses received an above average number of “N/R” assessments. Average values for each variable are presented in **Table 3**. While comparative assessment of studies is not the primary focus of our analysis and the BMJ checklist is not intended to produce a quantitative assessment, the distribution of values and the average value for each article is presented in **Appendix 6** and **Appendix 7**.

**Table 3:** BMJ Checklist Values across all Items.

| **Table 3: BMJ Checklist Values across all Items** |  |  |  |  |  |  |
| --- | --- | --- | --- | --- | --- | --- |
| **BMJ Checklist Item** | **Total 2s** | **Total 1s** | **Total 0s** | **Total N/Rs** | **Total N/As** | **Average Value\*** |
| The research question is stated | 47 | 0 | 0 | 0 | 0 | 2.00 |
| The economic importance of the research question is stated | 25 | 13 | 9 | 0 | 0 | 1.34 |
| The viewpoint(s) of the analysis are clearly stated and | 31 | 11 | 5 | 0 | 0 | 1.55 |
| justified |  |  |  |  |  |  |
| The rationale for choosing the alternative programs or interventions compared is stated | 44 | 3 | 0 | 0 | 0 | 1.94 |
| The alternatives being compared are clearly described | 42 | 5 | 0 | 0 | 0 | 1.89 |
| The form of economic evaluation used is stated | 39 | 3 | 0 | 5 | 0 | 1.93 |
| The choice of form of economic evaluation is justified in relation to the questions addressed | 43 | 0 | 0 | 4 | 0 | 2.00 |
| The source(s) of effectiveness estimates used are stated | 43 | 1 | 0 | 1 | 2 | 1.98 |
| Details of the design and results of effectiveness study are given (if based on a single study) | 16 | 2 | 0 | 28 | 1 | 1.89 |
| Details of the method of synthesis or meta-analysis of estimates are given (if based on an overview of a number of effectiveness studies) | 10 | 10 | 6 | 20 | 1 | 1.15 |
| The primary outcome measure(s) for the economic evaluation are clearly stated | 46 | 1 | 0 | 0 | 0 | 1.98 |
| Methods to value health states and other benefits are stated | 13 | 2 | 13 | 11 | 8 | 1.00 |
| Details of the subjects from whom valuations were obtained are given | 14 | 5 | 11 | 10 | 7 | 1.10 |
| Productivity changes (if included) are reported separately | 1 | 0 | 4 | 42 | 0 | 0.40 |
| The relevance of productivity changes to the study question is discussed | 1 | 2 | 2 | 42 | 0 | 0.80 |
| Quantities of resources are reported separately from their unit costs | 15 | 9 | 21 | 0 | 2 | 0.87 |
| Methods for the estimation of quantities and unit costs are described | 26 | 12 | 4 | 0 | 5 | 1.52 |
| Currency and price data are recorded (year, currency of costs, break into key components) | 35 | 7 | 4 | 0 | 1 | 1.67 |
| Details of currency of price adjustments for inflation or currency conversion are given | 25 | 3 | 17 | 1 | 1 | 1.18 |
| Details of any model used are given | 38 | 4 | 1 | 1 | 3 | 1.86 |
| The choice of model used and the key parameters on which it is based are justified | 40 | 3 | 0 | 2 | 2 | 1.93 |
| Time horizon of costs and benefits is stated | 36 | 6 | 2 | 3 | 0 | 1.77 |
| The discount rate(s) is stated | 42 | 1 | 1 | 3 | 0 | 1.93 |
| The choice of rate(s) is justified | 22 | 4 | 18 | 3 | 0 | 1.09 |
| An explanation is given if costs or benefits are not discounted | 0 | 0 | 2 | 45 | 0 | 0.00 |
| Details of statistical tests and confidence intervals are | 8 | 1 | 32 | 5 | 1 | 0.41 |
| given for stochastic data |  |  |  |  |  |  |
| The approach to sensitivity analysis is given | 34 | 8 | 1 | 4 | 0 | 1.77 |
| The choice of variables for sensitivity analysis is justified | 25 | 5 | 11 | 5 | 1 | 1.34 |
| The ranges over which the variables are varied are stated | 36 | 2 | 3 | 5 | 1 | 1.80 |
| Relevant alternatives are compared | 38 | 3 | 5 | 1 | 0 | 1.72 |
| Incremental analysis is reported | 43 | 0 | 1 | 3 | 0 | 1.95 |
| Major outcomes are presented in a disaggregated as well as aggregated form | 43 | 0 | 1 | 3 | 0 | 1.95 |
| The answer to the study question is given | 47 | 0 | 0 | 0 | 0 | 2.00 |
| Conclusions follow from the data reported | 47 | 0 | 0 | 0 | 0 | 2.00 |
| Conclusions are accompanied by the appropriate caveats | 41 | 6 | 0 | 0 | 0 | 1.87 |

*\*Average quality values were calculated for each question by summing the 1s and 2s each article received across all studies then dividing that sum by the number of items for*
*which 0s, 1s, and 2s were possible*

### Assessment of Key Methodological Constructs

#### Perspective, scope, and parameter selection

For studies that based effectiveness estimates for preventive interventions on several sources (n=25), roughly a third (n=7) presented a thorough evidence synthesis, which outlined how they identified the parameters used in their analysis. A systematic literature review was conducted and a formal meta-analysis was completed for important variables in only four articles (**Appendix Table 5**).^29–32^

Fourteen articles either conducted micro-costing or referenced prior micro-costing analyses, while the rest opted for a macro-costing approach. Furthermore, several studies (n=7) adopted costing information from other, similar cost-effective analyses without justifying the primary source of the costing data.

Of studies with a clearly stated perspective, all presented at least a healthcare payer or healthcare system perspective. Three articles also included components of a societal perspective; two of these studies incorporated lost labor productivity costs into overall costs and one conducted two separate analyses from either the healthcare sector or societal perspective. No studies incorporated non-medical benefits of genetic screening or testing, such as the personal utility of non-actionable genetic information or psychological benefits of negative test results. Studies that only examined carrier screening were excluded from the review, though two studies either incorporated costs associated with assisted reproductive technology use by parents after a child’s genetic diagnosis or DALYs averted by decisions to avoid having children with genetic disease.^33, 34^ One study included a discussion of the privacy implications of familial cascade testing,^35^ although privacy costs were not incorporated into their model.

No studies calibrate their model using real-world data. Two articles attempted some form of internal or external model validation, although this was not done to inform model parameterization but rather to confirm that model outcomes aligned with data used within the model and external values (e.g., known prevalence of disease).^32, 36^

#### Use of Sensitivity/Uncertainty Analyses

While all evaluations considered in this review conducted sensitivity analyses, the depth, breadth, and presentation of analyses varied widely. The majority of studies (n=33) conducted some form of one-way or two-way deterministic sensitivity analyses and 19 of such studies presented the results in the form of a tornado diagram. Among the 29 studies that included a probabilistic sensitivity analysis (PSA), nine displayed PSA results in ICER scatter plots, 23 presented cost-effectiveness acceptability curves, but only three presented uncertainty intervals for primary estimates. Twenty-one studies conducted at least one scenario analysis and eight studies conducted at least one threshold analysis. Only two value of information analyses were conducted, which included an expected value of perfect information analysis and an expected value of partial perfect information analysis for specific parameter groups (e.g., treatment costs, probability of cancer recurrence) (**Table 2**).

#### Reporting Transparency

Both the study question and answer to the study question were clear in all papers. Only two studies did not clearly report the discount rate for their analysis, though many studies (n=19) did not provide a proper justification of why their specific rate was selected. Similarly, most papers (n=39) clearly articulated the year and price information of their cost units but only about half (n=24) reported whether or how these prices had been adjusted for inflation or currency conversion.

All articles presented both disaggregated outcomes (such as total QALYs gained or total healthcare costs) as well as final ICER calculations. However, less than half of the studies (n=23) based the population size on a real-world population. Only one article disaggregated intervention costs into specific categories unique to genetic screening and nine studies disaggregated costing results based on the generic source of costs such as genetic sequencing, disease prevention, and disease treatment.

For studies that reported results in the form of either QALYs or DALYs (n=37), about half (n=16) presented the valuation method or study by which their utility values were generated and slightly more than half (n=19) reported the population from whom these values were generated.

Of the 19 studies that reported the results of one-way sensitivity analyses in the form of a tornado diagram, 9 had figures that did not indicate the direction of the associations between each parameter and the ICER. It was also unclear for several studies (n=10) why certain variables were ultimately selected to be included in deterministic sensitivity analyses (such as tornado diagrams) and not others. While more than half of the studies conducted a probabilistic uncertainty analysis.

## DISCUSSION

### Overview of Major Findings

This systematic review analyzed the methodological quality of 47 recent economic evaluations of genetic screening or testing for monogenic disorders across disease arenas. There was substantial variation in model sophistication and reporting quality. Most articles satisfied basic criteria for their presentation of parameter values, model design, and results as well as their implementation and interpretation of sensitivity/uncertainty analyses. A few studies achieved higher levels of sophistication or quality and can serve as exemplars for future work.^32, 34, 37–41^

### Recommendations for Future Evaluations and Exemplar Cases

While uniformity of evaluation design and reporting should not come at the cost of analytic flexibility, the heterogeneity of quality assessed in our review suggests the importance of further training to develop high-quality economic evaluations of genetic screening/testing. Scholars are encouraged to reference one or more of the guidelines that have been published over the past 20 years; these guidelines demonstrate near-consensus on the key elements of an economic evaluation.^19, 42^ Within the last five years, several textbooks have also been published on the proper methodological approach to economic evaluation.^3, 7, 28^

Informed by our assessment and considering authoritative sources, we make several recommendations for future economic evaluations of genetic testing/screening. Our recommendations focus on three arenas that consistently caused difficulty for articles considered in our review (parameter selection, use of sensitivity/uncertainty analyses, and reporting transparency). **Table 4** summarizes this discussion along with several exemplar cases from our review are provided to demonstrate recommended practices.

**Table 4:** Review-Informed Recommendations across Methodological Constructs.

| *Methodological Construct* | *Identified Challenge* | *Emphasized Recommendation* | *Exemplar Studies Identified in Systematic Review* |
| --- | --- | --- | --- |
| Perspective, scope, and parameter selection | As the variety of genetic testing/screening interventions expand (e.g., full-gene sequencing, multi-gene panels, whole exome or genome sequencing), it is difficult to track the accuracy of these interventions (e.g., sensitivity and specificity) | For parameter values that are especially influential or uncertain, conduct systematic reviews (with or without meta-analyses, depending on the consensus of the review); provide justifications for variations in parameter values when consensus is not available. For parameter values that are likely to change in different environments, base estimates on available evidence and justify choices. | In the context of familial hypercholesteremia, Crosland and colleagues<sup>43</sup> conducted a systematic review to determine the diagnostic accuracy of the Simon Broome and Dutch Lipid Clinic Network clinical assessment tools (incidentally, the review also determined the absence of information available to inform uptake probabilities)<br>See also: McKay<sup>44</sup> and Asphaug<sup>32434432</sup> |
|  | Estimating the costs of implementing a new genetic screening or testing intervention in practice, or the ongoing costs such as training or clinical decision support systems that need to be maintained over time to support intervention | Conduct micro-costing to estimate the varied sources of cost and categories of cost within the intervention, especially for analyses centered on changes in the way resources are delivered within a specific program or diagnostic odyssey | Asphaug and colleagues<sup>32</sup> used a departmental micro-costing analysis to estimate the cost of materials and equipment as well as direct labor, indirect labor, overhead, capital, and maintenance services for all scenarios included in the model.<br><sup>69</sup>See also: Crosland 2018<sup>43</sup>; Snowsill 2015<sup>37</sup> and 2017<sup>38</sup>; Compagni 2013<sup>91</sup>; Vrijenhoek 2018<sup>94</sup> |
|  | When implementing genetic analyses, cost-effectiveness may not be clear for all stakeholder perspectives. It is challenging to appropriately capturing all potential benefits from genetic analyses (e.g., secondary findings or non-health-related personal or reproductive utility) across these perspectives including distinguishing benefits from screening and cascade testing | To ensure all relevant impacts of the intervention have been considered from all appropriate perspectives (e.g., healthcare and societal as distinct), refer to the “Impact Inventory” (developed by the Second Panel on Cost-Effectiveness in Health and Medicine) | Lázaro and colleagues<sup>46</sup> demonstrated that family cascade testing was shown to be cost-effective (i.e., compared to usual care, the additional cost of testing was considered worthwhile given the additional benefits brought) when using the healthcare sector perspective and dominant (i.e., screening was both less costly and more effective than usual care) when using the societal perspective, primarily due to the days off work that testing prevented.<br>See also: <sup>46</sup>Asphaug 2019<sup>32</sup> |
| Use of Sensitivity/Uncertainty Analyses | The cost of genetic screening and testing interventions is constantly being updated | Conduct threshold analyses to interrogate key parameters that may change in response to policy decisions, | Naylor and colleagues<sup>53</sup> conducted a threshold analysis to predict the minimum prevalence of pathogenic variants for maturity-onset diabetes of the |
|  | (often becoming cheaper), which has dynamic implications for the cost-effectiveness of such interventions | programmatic design, or other exogenous factors, such as the cost of genetic screening necessary for an intervention to be cost-effective | young (MODY) at which screening would become cost-saving. Rubio-Terrés and colleagues<sup>55</sup> find that the cost of the new genetic tool Thrombo inCode® would need to fall substantially for it to be cost-effectively used to screen for risk of venous thromboembolism in Spain.<br>See also: Kwon 2019<sup>54</sup> |
|  | Appropriately accounting for potential uncertainty of information, such as the population prevalence of genetic variants or variants of unknown significance | Conduct value of information analyses to quantify the value of investing in research activities that generate additional evidence that lessens parameter uncertainty | Asphaug and colleagues<sup>32</sup> conducted an expected value of partial perfect information (EVPPI) for select parameter groups (including relative cancer risk, pathogenic variant prevalence, cost of cancer treatment, utility weights), which estimated the net monetary benefit from the removal of uncertainty around parameter values. The authors determined that gaining certainty about the relative cancer risk associated with specific pathogenic variants and the cost of breast cancer treatment had the highest per person EVPPI. This analysis prompted the authors to advocate for variant-specific prevalence data, which would allow for within-gene stratification in models.<sup>32</sup> |
|  | Genetic testing/screening interventions may be improved by several adaptations to the screening algorithm (e.g., which sub-populations to target) or investments in outreach (e.g., additional assistance to contact relatives of index cases), for which the cost-effectiveness is unclear and will need to be studied further | Conduct scenario analyses to learn about the relationship between such choices and estimated incremental cost-effectiveness | For instance, Gansen and colleagues<sup>35</sup> used scenario analyses to consider whether intensified outreach for cascade testing is cost-effective. For a detailed description of how scenario analyses were used across Familial Hypercholesterolemia studies, see **Appendix 8**<br>See also: <sup>35</sup>Snowsill 2015<sup>37</sup> and 2017<sup>38</sup>; McKay 2018<sup>44</sup>; Chen 2015<sup>47</sup> |
| Reporting Transparency | Genetic testing/screening interventions may lead to non-health-related changes in utility resulting from new awareness of having a | Identify valuation studies (i.e., studies attempting to assess the utility of distinct health states) among those with your genetic condition of interest, or those that closely parallel that | When presenting the utility values selected for individuals with breast or ovarian cancer, Müller and colleagues<sup>56</sup> clearly articulated the populations in which valuation studies were conducted (women with a present pathogenic variants/breast cancer or |
|  | condition, or health-related changes in utility not commonly described in the literature | condition; clearly articulate the target populations in which and valuations methods by which the studies were conducted to derive health state utility values | women from a healthy reference group), the valuation methods used across different studies (time trade-off [TTO] or standard gamble [SG]), and the reason for ultimately preferring one set of studies over another (SG more accurately reflected health-related quality of life compared to TTO, per their analysis).<sup>56</sup> |
|  | The costs of genetic testing/screening programs are constantly evolving, often at a different pace than other medical goods | Specify inflation or cost adjustments to medical commodities that have increased or decreased in price relative to the rest of the industry | Gansen and colleagues<sup>35</sup> identified medical costs that had been updated and how they were updated (using consumer price indices and purchasing power parity) since a publication of results using the same model four years prior, including the impact of new classification of tests relevant to Lynch Syndrome (though the specific classification was not mentioned).<sup>35</sup> |
|  | Genetic testing/screening interventions are often composed of several distinct activities which all demand varying resources costs, such as genetic counseling and clinical genetics, phlebotomy and ordering, and sequencing, analysis, and interpretation | When modeling and reporting the costs of the interventions, disaggregate intervention costs into specific categories unique to the genetic condition or disaggregate generic sources of cost into relevant categories for the testing or screening program | Ademi and colleagues<sup>40</sup> helpfully disaggregated intervention costs into specific categories unique to the genetic condition: “disease costs”, “intervention costs” and “screening and imaging” (although the specific item costs attributed to each category is not clear); if genetic testing/screening costs were to substantially change following their publication, readers would be more able to account for those changes and recalculate cost-effectiveness outcomes, thereby preserving the value of the original evaluation.<br>See also: <sup>40</sup>Leenen 2016<sup>57</sup>; Eccleston 2017<sup>58</sup>; Hoskins 2019<sup>59</sup> |

#### Perspective, Scope, and Parameter Selection

A central challenge in conducting any economic evaluation is employing expert judgement and the evidence synthesis needed to select or estimate parameter values for the model. A formal systematic review with or without meta-analysis should be attempted for parameter values that are especially influential, uncertain, or likely to change in different environments (e.g. as a consequence of policy decisions).^3434432^

Most economic evaluations of genetic testing/screening take a simplistic view of genetic analysis costs, often ignoring costs of implementation and patent outreach. For more realistic integration of the costs incurred by genetic testing/screening, micro-costing is recommended.^7, 45^ Micro-costing is especially important for analyses centered on changes in the way resources are delivered within a specific program or diagnostic odyssey, which is often the case for innovative genetic medicine programs.^28^ ^32^Micro-costing may not be suitable for studies primarily concerned with nationally aggregated or long-run costs, and the importance of various sub- components may depend on the perspective.

Economic evaluations of genetic testing/screening should evaluate value across relevant stakeholders, including but not limited to payer and societal perspectives. Genetic analyses are unusual in that they has implications not just for the individual being tested but also for family members, who may or may not be covered by the same payer. For settings without a single-payer, including family members in models requires careful consideration of how and even whether cascade testing is relevant in a payer-perspective analysis. Moreover, it has been strongly recommended that economic evaluations report *two* standard reference case perspectives: one from the healthcare payer perspective (i.e. formal healthcare sector costs borne by third-party payers or paid for out-of-pocket by patients) and, in parallel, one from the societal perspective (i.e. including patient/family time costs involved in receiving an intervention and for self-management).^3^ Presenting a reference case from a particular third-party payer (e.g. the federal government, a single healthcare system, or a particular insurance company) can also be warranted, though care should be taken to consider whether the covered population is stable, especially when benefits may lag many years behind initial investments (e.g., crossing Medicaid and Medicare programs or attrition from insurance plans). Presenting both analytical perspectives in tandem clarifies how value may vary substantially among key stakeholders. ^46^

To account for the balance between the burden of screening and recovered productivity, future studies should refer to the “Impact Inventory” developed by the Second Panel on Cost-Effectiveness in Health and Medicine to guide which costs should be considered from either perspective.^5^ This resource was used by only one study in this review.^32^ For the specific context of genomic screening programs, Fragoulakis and colleagues outline several direct costs (e.g., healthcare payer costs) and indirect costs (e.g., patient productivity lost and family expenses) which may also serve as a useful guide. Examples of indirect costs modeled in reviewed studies included work productivity lost because of illness^46^ and physician visits.^47^ Screening for genetic conditions in the general population may also lead to non-health-related changes in utility resulting from new awareness of having a condition that either requires additional interaction with the health care system or cannot be addressed medically.

Model calibration is a process used in economic evaluations to improve the accuracy of parameters that cannot be directly measured, leveraging available data that can be matched with the model.^48^ Calibration efficiently searches the space of plausible parameter values to find those which optimize the model’s fit to real-world data.^49^ Calibration is not always necessary but should be used when it can reduce the amount of parameter uncertainty in the model, especially for the most influential or actionable model parameters.^50^

Authors should pay special attention to test performance in the model. True clinical sensitivity is extremely difficult to measure for most conditions, and categories of possible test results vary between diagnostic testing, family cascade testing, and population screening.^51^ The probability of further interaction between the healthcare system and patient will differ based on how these categories are reported. Evaluators should ensure that their modeling of test results accurately reflects both what is known about the clinical sensitivity and specificity of the genetic test and how that knowledge is translated into diagnostic protocols, which may vary across sites of implementation.

### Use of Sensitivity/Uncertainty Analyses

Beyond reporting outcomes of a base case, economic evaluations of genetic testing/screening should identify and consider the impact of stochastic, parameter, and/or structural uncertainty as well as patient heterogeneity. Analyses should distinguish between variability in inputs that may affect outcomes (sensitivity analysis) and uncertainty in model inputs that may alter the uncertainty of model conclusions (uncertainty analysis).^28, 52^ As with all economic evaluations, it is challenging to estimate the collective impact that uncertainty within individual parameters will have on the uncertainty of overall model outcomes. We strongly recommend studies to conduct a ***Probabilistic Uncertainty Analysis (PUA)*** and to use the PUA results to clearly report the degree of uncertainty of estimates for primary outcomes of interest (i.e., confidence intervals). Given their likely dramatic impact on model outcomes, we recommend studies to consider the following parameters within their PUA: pathogenic variant prevalence (which depends on the target population and clinical scenario), probability of referral to genetic counseling and genetic testing uptake, likelihood of clinical outcomes (based on penetrance and expressivity of the condition), uptake/adherence and efficacy of interventions in symptomatic and pre-symptomatic individuals, morbidity and mortality in the absence of a genetic diagnosis, and cost of genetic analysis, implementation of interventions, and care used as part of post-result clinical interventions.

Studies should incorporate ***threshold analyses*** (a type of sensitivity analysis) to interrogate key parameters that may change in response to policy decisions, programmatic design, or other exogenous factors. Threshold analyses identify the minimum or maximum value for a given parameter that results in the intervention meeting willingness-to-pay thresholds. In the context of genetic testing/screening, this may fruitfully be applied to parameters such as the prevalence of the pathogenic variant being screened, with the assumption that programs could be developed to target populations with a critical prevalence rate (e.g., those with a clinical history suggestive of genetic disease). Threshold analyses could also determine the minimum rate of uptake for accepting genetic testing/screening or prophylactic interventions for screening to become cost-effective.^5354^ It is widely appreciated that genetic laboratory costs have fallen over the past decade, and there is speculation over whether testing prices will continue to fall or may even increase if testing companies capture greater control of markets. This value should also be strongly considered for threshold analyses. ^55^

As with any novel intervention, many parameters necessary to evaluate genetic testing/screening are fixed but unknown or uncertain. Some of these parameters, such as the prevalence of pathogenic variants in populations, could be studied using biobank or cohort studies and epidemiological research methods. ***Value of information analyses*** should be conducted to quantify the value of investing in research activities that generate additional evidence that lessens parameter uncertainty.^3^ This type of analysis informs what research is most valuable—essential information for researchers and funders.

***Scenario analyses*** should be used to estimate structural uncertainty or to compare different intervention approaches in a model. In the context of genetic testing/screening, they could be used to consider alternative scenarios in which more energy is dedicated to certain sub- populations or the diagnostic pathway is slightly modified for these sub-populations. ^3537,38^The consistent incorporation of scenario analyses will not only make models more informative (by calling attention to particularly uncertain or variable parameters) but also improve methodological rigor as authors are forced to critically think about the specific questions that their model must be designed to address.

### Reporting Transparency

The amount of content necessary to properly present an economic evaluation is often too much to fit in a single manuscript, prompting evaluators to reference secondary literature.^21^ When referencing secondary literature (especially for parameter estimation), summary information should be available within the main manuscript or appendix for readers to understand the context and methods behind the results produced from that literature. ^56^

When price transformations are necessary—either between different years or between different currencies—authors must be clear what year was used as the benchmark and what exchange rate was employed for the transformation. When adjusting for inflation, authors should use inflation rates unique to the medical industry.^28^ When relevant, inflation or cost adjustments should be specific to medical commodities that have increased or decreased in price relative to the rest of the industry (e.g., when a patented drug becomes available in generic forms).^35^Evaluators should also clearly identify when monetary amounts included in the model reflect price or cost estimates; we recommend accounting for all the associated costs of a medical good or activity.^28^ The cost of the same genetic analysis may also vary considerably depending on the equipment used, throughput level, and sequencing method; we recommend clearly identifying the sub-components of costs associated with the genetic analysis.

We strongly recommend disaggregating the outcome of cost-effectiveness analyses into total costs and total effectiveness. Disaggregation is especially useful if the size of the model population is reported and corresponds to a real-world population rate. This allows for population-wide health and economic impacts (e.g., a budgetary impact analysis) to be reported in addition to per-person cost and effectiveness. When expressing the total costs associated with any screening strategy, it is also recommended that authors report both total costs as well as costs disaggregated into relevant categories. This categorization provides a clear depiction of which aspect of genetic testing/screening is responsible for incremental cost differences. Detecting sources of incremental variation is especially important for a field such as genetics in which materials and activities are rapidly changing costs.^4056–59^

### Alignment with Similar Systematic Reviews

Several recent systematic reviews of economic evaluations of genetic screening have been conducted for either specific populations or a more limited set of medical conditions. While prior reviews primarily covered older studies, were limited to specific genetic conditions, and were not as comprehensive as our own regarding methodological assessment, these reviews have identified many of the same limitations in economic evaluations our review has exposed. These include emphasizing the healthcare payer or health system costing perspectives over societal perspectives,^60–63^ dependence on macro-costing strategies and adopting costing estimates from other, similar studies,^64^ and limited or opaque use of complex sensitivity analyses.^60, 65^ The performance of our articles as measured by the BMJ checklist is also consistent with two recent reviews of economic evaluations of genetic testing that employed the BMJ checklist.^20, 65^ Both these reviews found that most studies failed to provide a rigorous description of how costs were derived, provided no description for how disparate sources were synthesized to establish effectiveness estimates, failed to appropriately adjust price or currency or report such adjustment, and had limited description of the valuation methods by which utility weights were calculated or characterizations of the population from which they were derived.

### Study Limitations

There are several limitations to this systematic review. Firstly, our assessment mechanism gave equal weight to all items, implying that all items were of equal ease to achieve and of equal importance to the methodological quality of an article when important inequalities likely exist across both dimensions. To account for this limitation, we have focused our discussion on those items which we believe to be of greater importance to overall quality and have provided recommendations to facilitate ease of achievement. Secondly, this review does not consider the influence methodological limitations may have on the primary or secondary outcomes of studies. For instance, an opaque presentation of parameter derivation may complicate a reader’s ability to interrogate the integrity of a model, though these parameters may ultimately be the most appropriate leaving results unbiased. On the other hand, the lack of a PSA may indirectly hide the fact that primary outcomes are widely variable and cannot be interpreted with high confidence. Future research should consider which methodological features of an article may have the largest influence on outcomes. Thirdly, there is an abundance of methodological detail that went beyond the scope of this review, such as how well the structure of the model reflected the actual decision nodes within the healthcare system under study and whether a comprehensive selection of alternative strategies was considered for each model. This level of granularity is best suited for reviews with a much more limited scope than the one we conducted.

## Conclusion

Economic evaluation of genetic medicine has been recently accelerating. Our review considered the methodological quality of such studies and demonstrated that, with notable exceptions, many studies fell short across several key methodological criteria. Improvements in these arenas highlighted above would enhance the extent to which outcomes can be understood, translated, and faithfully replicated. Renewed attention to the methodological design of future economic evaluations of genetic testing/screening is warranted. Future economic evaluations in this space should adhere to established guidelines and may benefit from considering the specific recommendations and exemplar articles identified in this review.

## Data availability

All articles included in this review are accessible online, and the search terms used to query these articles can be found in the Appendix.

## Data Availability

Data used to create this review is available upon request

## Acknowledgments

We wish to acknowledge Dr. Gail Henderson for both her help with the design and scope of this systematic review as well as for funding early research assistant support through the Center for Genomics and Society at UNC (grant ID: 2P50 HG004488), a National Human Genome Research Institute (NHGRI)-funded center. We also wish to acknowledge Hailey James for developing our query search strategy, contributing to study planning, and conducting article screening. This study was partially supported by another grant from the support from the NHGRI at the National Institutes of Health (grant ID: 2U01 HG006487).

## Author Information

Conceptualization: K.H.L., K.S., I.G., K.J., J.S.B., K.H.; Data curation: K.S., I.G.; Formal analysis: K.J., K.S., K.H.L.; Funding acquisition: K.H.L., J.S.B.; Investigation: K.J., K.S., I.G., K.H., K.H.L.; Project administration: K.J., K.S.; Supervision: J.B., K.H.L.; Writing—original draft: K.J., K.S., K.H.L.; Writing—review and editing: I.G., K.H., J.S.B., K.J., K.S., K.H.L.

## Ethics Declaration

This study was determined to be non-human subjects research by the University of North Carolina IRB.

