## Appendix for "A Systematic Review of the Methodological Quality of Economic Evaluations in Genetic Screening and Testing for Monogenic Disorders"

**Appendix 1: Distinguishing between Genetic *Testing* and Genetic *Screening***

Within genetic and genomic medicine, the same laboratory analysis might be used in different clinical scenarios, distinguished largely by the pre-test probability of a clinically relevant genetic variant being present in the individual. Although there are no strict borders or defined parameters to characterize a given genetic analysis as “testing” versus “screening,” for the purposes of this work we will use “genetic testing” when referring to a clinical diagnostic setting in which a patient is symptomatic of a possible/presumed/definite Mendelian condition or at risk for that condition due to a strong family history, and the genetic analysis is being done to confirm/rule-out a specific diagnosis or to evaluate a differential diagnosis containing several or many possible diagnoses. Conversely, we use “genetic screening” when the individual being tested is not symptomatic of a Mendelian condition and not otherwise at greatly increased risk due to family history (i.e., the individual is close to the general population risk) but genetic analysis is being done to establish the presence of a molecular diagnosis that indicates risk to develop symptoms of the condition. Cascade testing for a known familial variant in a person who does not yet have symptoms could be considered a form of diagnostic testing because the risk to have that variant can be as high as 50%. Secondary findings identified during the course of a broad genomic analysis such as exome or genome sequencing can be thought of as a type of “opportunistic screening” in that variants in genes that are considered “clinically actionable” may be found in an individual before symptoms related to those conditions manifest. Although cascade testing of family members for a known pathogenic variant, diagnostic testing of at-risk but asymptomatic individuals based on a strong family history of disease, and analysis of clinically actionable secondary findings may have different pre-test probabilities of genetic variants being identified in a given population, they have similarities with general population screening in terms of subsequent downstream interventions being targeted at prevention of symptoms that may develop in the future, as opposed to interventions that are intended to treat an existing disease.  We think that it can be useful to consider these different clinical scenarios when performing economic evaluations, in order to differentiate between measures of the direct impact of the genetic testing on health utilization and health outcomes in affected individuals versus the impact of preventive interventions (which may require modeling the impact of false positive results as well as overdiagnosis due to variable expressivity and incomplete penetrance of genetic conditions).

**Appendix 2: Search Strategy**

**Search Databases and Timeline**

Pubmed, Cochrane, Embase, and Web of Science were queried for studies that had terms related to both genetic testing and economic evaluation. The search was limited to full text, peer-reviewed, English language studies published since 1/1/2013, with the most recent search conducted on 9/10/2019. Covidence systematic review software (Veritas Health Innovation, Melbourne, Australia. Available at [www.covidence.org](http://www.covidence.org/)) was used for title/abstract and full-text review.

**Search Terms**

Pubmed:

((("Genetic Testing"[Mesh] AND "Cost-Benefit Analysis"[Mesh]) OR ("Genetic Testing/economics"[Mesh]) OR ("Genetic Carrier Screening"[Mesh] AND "Cost-Benefit Analysis"[Mesh])) OR (("mutation test" OR "mutation tests" OR "mutation testing" OR "mutation testings" OR "mutation screen" OR "mutation screening" OR "mutation screens" OR "mutation screenings" OR "genetic test" OR "genetic testing" OR "genetic tests" OR "genetic testings" OR genetic screen* OR "personalized medicine" OR population test* OR population screen* OR genomic diagnostic test* OR "next-generation sequencing" OR "whole-exome sequencing" OR "whole exome sequencing" OR "whole genome sequencing") AND ("cost effective" OR "cost-effective" OR "cost-effectiveness analysis" OR "cost effectiveness" OR "cost-effectiveness" OR "economic evaluation" OR "cost-utility" OR "cost utility" OR "cost-benefit" OR cost benefit*))) NOT (animals [mh] NOT (humans [mh] AND animals [mh])) AND ( "2013/01/01"[PDat] : "3000/12/31"[PDat])

Web of Science:

TOPIC: ("mutation test*" OR "mutation screen*" OR "genetic test*" OR "genetic screen*" OR "carrier screen*" OR "personalized medicine" OR "population test*" OR "population screen*" OR "genomic diagnostic test*" OR "next-generation sequencing" OR "whole-exome sequencing" OR "whole exome sequencing" OR "whole-genome sequencing" OR "whole genome sequencing") AND ("cost-effective*" OR "cost effective*" OR "economic evaluation" OR "cost-utility" OR "cost utility" OR "cost-benefit" OR "cost benefit")

Refined by: DOCUMENT TYPES: ( ARTICLE ) Refined by: English

Timespan: 2013NA019. Indexes: SCI-EXPANDED, SSCI, A&HCI, CPCI-S, CPCI-SSH, BKCI-S, BKCI-SSH, ESCI, CCR-EXPANDED, IC.

Embase:

('mutation test/exp' OR 'mutation screen/exp' OR 'genetic test'/exp OR 'genetic screen'/exp OR 'carrier screen'/exp OR 'personalized medicine' OR 'population test'/exp OR 'population screen'/exp OR 'genomic diagnostic test'/exp OR 'next-generation sequencing' OR 'whole-exome sequencing' OR 'whole exome sequencing' OR 'whole-genome sequencing' OR 'whole genome sequencing' OR 'genetic screening'/de OR 'next-generation sequencing'/de OR 'whole exome sequencing'/de) AND ('cost-effective'/exp OR 'cost effective'/exp OR 'economic evaluation'/exp OR 'cost-utility' OR 'cost utility' OR 'cost-benefit' OR 'cost benefit' OR 'cost-effectiveness analysis'/de OR 'cost utility'/de OR 'cost benefit'/de) AND (2013:py OR 2014:py OR 2015:py OR 2016:py OR 2017:py OR 2018:py OR 2019:py) NOT [medline]/lim

Cochrane:

(([mh "genetic testing"] OR [mh "genetic carrier screening"] OR [mh "whole genome sequencing"] OR “mutation test*” OR “mutation screen*” OR "genetic test*" OR “genetic screen*” OR “carrier screen*” OR "personalized medicine" OR "population test*" OR "population screen*" OR "genomic diagnostic test*" OR "next-generation sequencing" OR "whole-exome sequencing" OR "whole exome sequencing" OR "whole genome sequencing" OR “whole-genome sequencing”) AND ([mh "cost-benefit analysis"] OR "cost-effective*" OR "cost effective*" OR "economic evaluation" OR cost-utility OR "cost utility" OR cost-benefit OR "cost benefit")) Limits: publication year 2013NA019, in Trials

**Review Process**

Two reviewers independently completed title and abstract review (KS, HJ, KH). At this stage, conflicts resulted in inclusion. Two independent reviewers completed full-text review, categorizing excluded studies by reason for exclusion (KS, HJ, KH). At the full-text review stage, conflicts (inclusion vs. exclusion or reason for exclusion) were resolved by consensus between the two reviewers. If conflicts remained, they were resolved at full team meetings. All included studies were confirmed by the full team (KS, HJ, KH, JB, KHL, GH).

| **Appendix Table 3: Codes and Summary Templates** | | |
| --- | --- | --- |
| **Code** | **Memo** | **Summary Template Format** |
| Study question |  | Is it cost-effective to screen POPULATION for TEST/GENE compared to REFERENT? |
| Type of evaluation\CEA | Definition: Cost effectiveness analysis -- outcomes are direct endpoints, like deaths averted. No summary required | [Type of Evaluation] |
| Type of evaluation\CUA | Definition: Cost utility analysis -- outcomes in QALYs or similar. Most common. No summary required | [Type of Evaluation] |
| Type of evaluation\CMA | Definition: Cost minimization analysis. No summary required. | [Type of Evaluation] |
| Type of evaluation\CBA | Definition: Cost benefit analysis -- outcomes are in monetary values ($$). No summary required | [Type of Evaluation] |
| Type of study\Simulation model | Definition: The study involved use of computer-generated simulation to model cost and impact data. No summary required | [Type of Study] |
| Type of study\Trial-based study | Definition: The study involved use of data generated by trials (i.e., clinical trials) to model cost and impact data. No summary required | [Type of Study] |
| Time horizon | Enter summary (follow exact formatting): lifetime, XX years, XX months, not stated | Lifetime. OR XX years. OR Age 100. |
| Time step | Enter summary (follow exact formatting) | XX weeks, XX months, XX years, not stated, no mid-cycle corrections |
| Perspective\Societal | Definition: Includes non-health sector costs. No summary required | [Perspective] |
| Perspective\Government | Definition: Includes costs to the government. No summary required | [Perspective] |
| Perspective\Healthcare sector | Definition: Includes all health related costs including patient out of pocket. No summary required | [Perspective] |
| Perspective\Healthcare sector plus time costs | Definition: Includes patient out of pocket and patient time costs. No summary required | [Perspective] |
| Perspective\Payer | Definition: Includes payer costs only. No summary required | [Perspective] |
| Perspective\Patient | Definition: Includes patient out of pocket costs and time costs only. No summary required | [Perspective] |
| Perspective\Not stated | Enter summary (follow exact formatting): not stated, appears to be XX perspective OR not stated, no clear perspective | [Perspective] |
| Baseline year of evaluation | Enter summary (follow exact formatting) | XXXX (e.g. 2012) |
| Discounting | Enter summary (follow exact formatting) | XX% costs, costs and outcomes, or outcomes (specify what values are discounted) |
| Model type\Decision tree | Definition: Can code as multiple. No summary required | [Model Type] |
| Model type\Markov model | Definition: Can code as multiple. No summary required | [Model Type] |
| Model type\Micro or individual simulation | Definition: Can code as multiple. No summary required | [Model Type] |
| Model type\Other | Enter summary (follow exact formatting): XXXX model | [Model Type] |
| Model type\Not stated | Enter summary (follow exact formatting): not stated, appears to be XX model OR not stated, no clear model | [Model Type] |
| Clinical condition\Cancer\Breast | Definition: Can code as multiple. BRCA maps to breast and ovarian. No summary required | [Clinical Condition] |
| Clinical condition\Cancer\Ovarian | Definition: Can code as multiple. BRCA maps to breast and ovarian. Lynch maps to endometrial, colorectal and ovarian. No summary required | [Clinical Condition] |
| Clinical condition\Cancer\Endometrial | Definition: Can code as multiple. Lynch maps to endometrial, ovarian, and colorectal. No summary required | [Clinical Condition] |
| Clinical condition\Cancer\Colorectal | Definition: Can code as multiple. Lynch maps to endometrial, ovarian, and colorectal. No summary required | [Clinical Condition] |
| Clinical condition\Cancer\Prostate | Definition: Can code as multiple. No summary required | [Clinical Condition] |
| Clinical condition\Cancer\Other | Definition: Can code as multiple. No summary required | [Clinical Condition] |
| Clinical condition\Cardiovascular\Hypertension | Definition: Can code as multiple. No summary required | [Clinical Condition] |
| Clinical condition\Cardiovascular\Cardiomyopathy | Definition: Can code as multiple. No summary required | [Clinical Condition] |
| Clinical condition\Cardiovascular\Hypercholesterolemia | Definition: Can code as multiple. No summary required | [Clinical Condition] |
| Clinical condition\Cardiovascular\Arrhythmia | Definition: Can code as multiple. Includes long QT syndrome. No summary required | [Clinical Condition] |
| Clinical condition\Endocrine\Diabetes | Definition: Can code as multiple. No summary required | [Clinical Condition] |
| Clinical condition\Neurological\Neurodevelopmental | Definition: Can code as multiple. No summary required | [Clinical Condition] |
| Clinical condition\Neurological\Neuromuscular | Definition: Can code as multiple. Includes intellectual disability, autism, seizures. No summary required | [Clinical Condition] |
| Clinical condition\Neurological\Neurodegenerative | Definition: Can code as multiple. No summary required | [Clinical Condition] |
| Clinical condition\Respiratory\AlphaNR antitripsin deficiency | Definition: Can code as multiple. No summary required | [Clinical Condition] |
| Clinical condition\Respiratory\Cystic fibrosis | Definition: Can code as multiple. No summary required | [Clinical Condition] |
| Clinical condition\Other\Smoking cessation | Definition: Can code as multiple. No summary required | [Clinical Condition] |
| Clinical condition\Other\Undiagnosed disorders | Enter summary (follow exact formatting): Undiagnosed disorder, XXXX (state undiagnosed disorder) | [Clinical Condition] |
| Clinical condition\Other\Hemochromatosis | Definition: Can code as multiple. No summary required | [Clinical Condition] |
| Clinical condition\Other\Telangiectasia | Definition: Can code as multiple. No summary required | [Clinical Condition] |
| Clinical condition\Other\Other | Enter summary (follow exact formatting) | [Clinical Condition] |
| Gene/Mutation(s) tested | Enter summary (follow exact formatting) | Targeted panel for [GENES] with del/dup analysis using MLPA, OR WES s |
| Sequencing method\WGS or WES | Enter summary (follow exact formatting) | [Diagnostic WGS, or Diagnostic WES] |
| Sequencing method\Targeted panel | Enter summary (follow exact formatting) | XXXX panel (e.g. 9-gene breast cancer panel, 57-gene ACMG panel) |
| Sequencing method\Single gene | Enter summary (follow exact formatting) | XXXX (e.g. Full gene sequence, or genotype) |
| Sequencing method\RNA sequencing | Enter summary (follow exact formatting) | XXXX (e.g. RNA sequencing of single gene product) |
| Referent | Enter summary (follow exact formatting) | XXXX (e.g. No testing) |
| Alternatives | Enter summary (follow exact formatting) | 1. Standard of care/referent.  2. 1st alternative/intervention.  3. 2nd alternative.  4. 3rd alternative. |
| Downstream from testing | Enter summary (follow exact formatting) | People who screen [positive/negative] will [activies downstream from testing] |
| Cascade testing | Enter summary (follow exact formatting) | Benefit from [cascade only] OR [proband and cascade] |
| Cascade testing\Benefit from cascade only | Definition: Cascade testing begins with the identification of an individual with the condition and/or a pathogenic variant associated with the condition and then extending genetic testing to his/her at-risk biological relatives. No summary required. | Benefit from cascade only |
| Cascade testing\Benefit from proband and cascade | Definition: A proband is an individual being studied or reported on. A proband is usually the first affected individual in a family who brings a genetic disorder to the attention of the medical community. No summary required. | Benefit from proband and cascade |
| Study population | Definition: Capture all study population attributes discussed (e.g. age (age of testing), sex, race/ethnicity, nationality (country/location), diagnosis, etc.). | [size of cohort] [Race/Nationality] [Sex] [Age] with [Clinical/Genetic Condition] (prevalence of condition = X%) |
| Cost units | Enter summary (follow exact formatting) | [Year] [Currency] |
| Source of costs | No summary required |  |
| Costing method |  | [Costing method] |
| Costing method\Gross Costing | Definition: Gross-costing is a cost estimation method that uses aggregated cost estimates for units of input and output that are large relative to the intervention being analyzed. No summary required. |  |
| Costing method\Micro-costing | Definition: Micro-costing is a cost estimation method that involves the direct enumeration and costing out of every input consumed in the treatment/intervention. No summary required. |  |
| Cost elements |  | Costs included [cost elements] |
| Cost elements\Disease prevention | The cost of any drug of prophylaxis medication to prevent the onset of future disease after being positively screened for genetic susceptibility. |  |
| Cost elements\Genetic test | Enter summary (follow exact formatting) | [cost elements] = cost |
| Cost elements\Clinical geneticist or counselor | Enter summary (follow exact formatting) | [cost elements] = cost |
| Cost elements\Disease management | The cost of treatment for the adverse event which is associated with the genetic variant |  |
| Cost elements\Time or opportunity costs | No summary required |  |
| Source of utilities | No summary required. |  |
| Outcomes\Non ICER Outcome | Definition: Code all non-ICER outcomes reported. Be careful to include units. Can code as multiple. Summarize briefly in own words. |  |
| Outcomes\ICER | Definition: Code all ICERs reported. Be careful to include referent group used in calculation and ICER units. Can code as multiple. Summarize in brief (your own words) | (Strategy # of alternative) compared to strategy (Strategy # of referent): [Currency Symbol] [VALUE]/QALY or [Currency Symbol][VALUE]/LYG.  **Include confidence interval in parentheses if reported |
| CE threshold | Enter summary (follow exact formatting): Enter willingness to pay or cost effectiveness threshold (e.g. 1.2 GDP per capita, $60,000) | [currency symbol] [value] / [unit of effectiveness] |
| Sensitivity analysis\One way or two way | No summary required. |  |
| Sensitivity analysis\Probabilistic | Enter summary (follow exact formatting) | Using a monte-carlo simulation (XX iterations), [Alternative] compared to [Referent] is cost-effective in [XX]% of simulated interations, at a threshold of XX/QALY, |
| Sensitivity analysis\3 most sensitive uncertain parameters | Enter summary: summarize each uncertain parameter in brief (your own words) | [Strategy X] compared to [Strategy Y]: (1) ______(+/-), (2) ________(+/-), (3) ________(+/-) |
| Sensitivity analysis\CE Plane | No summary required |  |
| Sensitivity analysis\Scatter Plot | No summary required |  |
| Sensitivity analysis\Threshold Analysis | Enter summary (follow exact formatting) | At a WTP of XX, [Alternative] will only be cost effective compared to [Referent] when [VAR] [Increases/Decrease] from [Old Level] to [New Level] |
| Sensitivity analysis\CEAC | No summary required |  |
| Sensitivity analysis\Scenario Analysis | No summary required |  |
| Sensitivity analysis\Tornado Diagram | No summary required |  |
| Value of information analysis | No summary required |  |
| Stated conclusion | Enter summary (follow exact formatting) | At a WTP threshold of XX, it IS/IS NOT cost-effective to screen POPULATION for TEST/GENE compared to REFERENT. |

**Appendix 4: BMJ Checklist Assessment Process and Rubric (Expanded Description)**

To assess the quality of papers considered in this review, we employed the 35 quality items outlined in the BMJ checklist. To aid the analysis of article quality, we developed an assessment system in which, for every article included in our review, each item on the BMJ checklist received a value which reflected how well the item was satisfied, roughly parallel to the original “yes”, “no”, “not clear”, and “not appropriate” responses available for the BMJ checklist. For each article in our review and each item on the BMJ checklist, we assigned a value of NA, NR, 0, 1 or 2 based on the following criteria. We assigned a value of 0 if the articles did not address the item whatsoever, 1 if the article addressed the item but in an incomplete way, and 2 if the article fully satisfied the item (with direct reference to the characteristics of that item as described in the BMJ checklist). A value of NA was given if the article had an appropriate response to the item, but the response was in the form of a citation to a document not directly affiliated with the study (i.e. this did not include the appendix or online supplement, which were both examined for each article as if part of the main manuscript). This would be the case, for instance, if an article did not explicitly describe the “Details of the subjects from whom valuations were obtained or given” within the main text of the manuscript but rather referenced the study in which the valuation was conducted. This value was created to distinguish between instances in which the response was nowhere to be found (which would receive 0 points) and instances in which the response was available in a separate but nevertheless referenced and accessible document (“secondary literature”). We did not want to outright penalize this delegation, as secondary literature sources are often established resources in the field. We rewarded studies with a value of 2 if the secondary literature was referenced with sufficient description to satisfy the quality item. We assigned a value of NR if the item was not applicable to the article’s study; for instance, a NR would be assigned to the variable “Productivity changes (if included) are reported separately” if studies did not include costs associated with productivity impacts. For items which could have been addressed in the main text and for which no external references were provided, we assigned a value of 0, 1 or 2. Prior to assessing all articles, the following assessment rubric was developed which articulated in detail what content constituted a NA, NR, 0, 1 or 2 for each item on the checklist

| **Appendix Table 4: BMJ Checklist Assessment Rubric** | | | | |
| --- | --- | --- | --- | --- |
| **Variable** | **Article contents needed to satisfy assessment value** | | | |
|  | **Value = NR** | **Value = 0** | **Value = 1** | **Value = 2** |
| The research question is stated |  | Research question is not stated | Research question is either not stated or not stated clearly in a way that considers both costs and outcomes. | BMJ: "the question should be phrased in a way that considers both costs and outcomes. The research question "Is drug X more costly than the existing therapy?" will provide incomplete information because the decision maker also needs to consider comparative effectiveness."  For a given perspective, sometimes the consequences are not health outcomes. They are the financial benefits of the health outcomes - not the health outcomes themselves (e.g., cost analysis of preventive cancer care from the payer perspective (Medicaid, etc.) where both cost and "outcome" [treatment or no cancer treatment required downstream] have been monetized). This is a full cost/consequence and should be given a 2. |
| The economic importance of the research question is stated |  | Economic important of the research question is not stated. | The research question is stated, but without any mention of the economic nature of the situation and why that nature prompts the question. | BMJ: "the question should be economically important (in terms of its resource implications) and be relevant to the choices facing the decision maker. The question "Is health promotion worthwhile?" does not meet this criterion because it fails to specify alternatives-worthwhile compared with what?" |
| The viewpoint(s) of the analysis are clearly stated and justified |  | The viewpoints of the analysis are not stated. | The viewpoint is merely stated, with no justification for why this viewpoint was chosen over others. | BMJ: "The research question should clearly state the viewpoint of the economic evaluation, and this should be justified. Possible viewpoints include those of the provider institution, the individual clinician or professional organization, the patient or patient group, the purchaser of health care (or third-party payer), and society itself. For example, hospital and other providers may need information to help in making procurement and related technology management decisions; individual clinicians to inform patient care decisions; health insurers or purchasers to support decisions on whether to pay for a procedure or which services to develop; and patients to know the level of costs they may incur in travelling to hospital or providing informal nursing care at home...Researchers should therefore identify key potential decision makers (government, purchaser, or provider) at the outset and be able to show that the research question posed will meet the needs of all key groups." This can be included in either the introduction or the methods section. |
| The rationale for choosing the alternative programs or interventions compared is stated |  | No rationale for choosing alternative programs or interventions is stated. | The decision of which alternatives will be compared is mentioned, but without any rationale for why those alternatives are the most salient to be compared to or with an inappropriate rationale. | BMJ: "The question "Is health promotion worthwhile?" does not meet this criterion because it fails to specify alternatives-worthwhile compared with what? Furthermore, any alternatives need to be realistic. An option of "doing nothing," or maintaining the status quo, should be included when appropriate...The choice of the alternative must be designed to help get as close a measure as possible of the opportunity cost of using the new treatment. In principle the comparator should be the most cost effective alternative intervention currently available. In practice the comparator is usually the most widely used alternative treatment. Unless current practice is "doing nothing," it is usually best not to use placebo as the comparator. Such a study could, however, if well conducted and reported, provide information for use in conjunction with studies of other treatments also compared with placebo." |
| The alternatives being compared are clearly described |  | The alternatives being compared are not listed. | The alternatives are merely listed, but without a full description of their mechanisms in such a way that a reader could reasonable "sketch-out" what is happening to patients in the other branches. | The description of alternatives is comprehensive--at least as much as is given for the reference case. One should be able to "sketch out" what is happening in all alternative cases. BMJ: "The alternatives being compared should be described in enough detail to enable the reader to relate the information on costs and outcomes to the alternative courses of action... Clear exposition of alternative treatment paths and the probabilities, cost, and outcomes linked to them should enable decision makers to use those parts of the analysis that are relevant to their viewpoint." |
| The form of economic evaluation used is stated |  | The form of economic evaluation used is not stated |  | The BMJ lists 4 options: Cost-effectiveness analysis, cost-utility analysis, cost-benefit analysis, cost-minimization analysis. If at least one of them is clearly stated, then the article receives a 2. |
| The choice of form of economic evaluation is justified in relation to the questions addressed |  | No justification is given for the form of economic evaluation used. | A justification is given for why this form was used as opposed to (or in conjunction with) the other three forms, but it is an inappropriate justification. | BMJ paraphrased: CBAs should be used to answer the question: "Is it worth achieving this goal?" or "How much more or how much less of society's resources should be allocated to pursuing this goal?" CEAs should be used to answer the question: "Given that a goal is to be achieved, what is the most efficient way of doing so?" or "What is the most efficient way of spending a given budget?". If the alternatives are equally effect and one just wants to compare their cost, a "cost-minimization" analysis is to be done. "The [final] category of evaluation, cost-utility analysis, lies somewhere between cost effectiveness and cost benefit analysis. It can be used to decide the best way of spending a given treatment budget or the health care budget." |
| The source(s) of effectiveness estimates used are stated |  | The source(s) of effectiveness estimates used are not stated |  | The source(s) of effectiveness estimates used are stated. |
| Details of the design and results of effectiveness study are given (if based on a single study) | Not based on a single study | No results of effectiveness study are clearly given. | Results of effectiveness study are given, but with no reference to the details/design of the study. | BMJ: "If the economic evaluation is based on a single effectiveness stud-for example, a clinical trial-details of the design and results of that study should be given-for example, selection of study population, method of allocation of subjects, whether analyzed by intention to treat or evaluable cohort, effect size with confidence intervals." |
| Details of the method of synthesis or meta-analysis of estimates are given (if based on an overview of a number of effectiveness studies) | Not based on a meta-analysis | No results of effectiveness study are clearly given. | Results of effectiveness studies are given, but with no reference to how those studies were synthesized. | BMJ: "If the economic evaluation is based on an overview of a number of effectiveness studies details should be given of the method of synthesis or meta-analysis of evidence-for example, search strategy, criteria for inclusion of studies in the overview." |
| The primary outcome measure(s) for the economic evaluation are clearly stated |  | The primary outcome measure(s) for the economic evaluation are not stated | The outcomes of the study are reported, but not until the results section of the paper (as opposed to where they should be introduced in the methods section), or they are never stated clearly. | Outcomes are described in the methods section of the paper and justification for why those outcomes were chosen should be given. BMJ: "The primary outcome measure(s) for the economic evaluation should be clearly stated-for example, cases detected, life years, quality adjusted life years (QALYs), willingness to pay...Authors using cost effectiveness analysis should explain why they have chosen a particular outcome measure for calculation of the ratio and reassure the reader that important outcomes are not being overlooked." |
| **Methods** to value health states and other benefits are stated | Health states have not been given values | The value of the health states (HRQoL) used are presented but without any description of the methods used to assign value to them. | Partial: Either the method or the population surveyed is given, but not both. | BMJ: "If health benefits have been valued details should be given of the methods used-for example, time trade off, standard gamble, contingent valuation-and the subjects from whom valuations were obtained-for example, patients, members of the general public, health care professionals." |
| Details of the subjects from whom valuations were obtained are given | Health states have not been given values | Valuations do not exist | Valuations exist but no details are given concerning the subjects from whom the valuations were obtained. | Valuations exist and details about the subjects from whom those valuations were obtained is stated. BMJ: "Values can be provided by the population at large or by a sample of patients with the condition for which the treatment is being evaluated. The choice depends on the perspective of the study. If the issue is allocating resources between competing programs the former might be used; if it is deciding the best way to treat a given condition the latter might be used. In reporting their results authors should explain why a particular source of values has been used." |
| Productivity changes (if included) are reported separately | Productivity values not provided | Productivity values referred to but never explicitly mentioned | Productivity values explicitly mentioned but are not separate from the main outcomes | BMJ: "Whatever estimation method is used, indirect benefits should be reported separately so that readers can decide whether or not they should be included in the overall result of the study." |
| The relevance of productivity changes to the study question is discussed | Productivity values not provided | relevance of productivity changes not discussed |  | Relevance of productivity changes (e.g., changes in labor time) is clearly discussed. |
| Quantities of resources are reported separately from their unit costs |  | Neither unit cost nor quantities of units are provided | Either just the unit cost or the quantity of the unit is provided | Both unit costs and the quantity of items is provided. BMJ: "Costing involves estimating the resources used-for example, days in hospital- and their prices (unit costs). These estimates must be reported separately to help the reader judge their relevance to his or her setting. When there are many cost items reporting should concentrate on the main costs." |
| Methods for the estimation of quantities and unit costs are described |  | Neither the methods nor the source for estimation of quantities of unit costs are not given | Just the source is given for estimation of unit costs | Source and the methods used in the source are given for the estimation of costs. That is, if a source is presented in a table, an explanation of the methodologies used in that source are presented in the manuscript. BMJ: "Outside the context of a trial, estimates of resource quantities should be based on data on real patients, collected either prospectively or retrospectively from medical records. The use of physician "expert panels" to estimate resource quantities, while common, runs the risk that respondents may give inaccurate estimates or specify the resources required for ideal care, rather than that provided in practice." |
| Currency and price data are recorded |  | Currency and price data not recorded. | Partially reported (e.g. total cost of hospitalization is given, but they do not break down into the cost/days OR they give the most important cost parameters but not all) | BMJ: "...the dates of both the estimates of resource quantities and prices should be recorded..." |
| Details of currency of price adjustments for inflation or currency conversion are given |  | Price and currency adjustments not given | Price and Currency adjustments are given but not done appropriately | Price and currency adjustments are given and are appropriately justified per the BMJ guidelines here. BMJ: "attention should be paid to the generalisation of cost estimates, since relative prices and the opportunities to redeploy resources may differ from place to place." Currency conversions should, when possible, be based on real purchasing power, rather than financial exchange rates, which fluctuate according to money market changes." |
| Details of any model used are given | If it's an RCT, single study ocst analysis (no modeling) | Model details not given | Model details are sparse or not transparent | Model details are thorough and transparent. BMJ: "Details should be given of any modelling used in the economic study-for example, decision tree model, epidemiology model, regression model...The key requirements are that the modelling should be explicit and clear. The authors should explain which of the reported variables/parameters have been modelled rather than directly observed in a particular sample; what additional variables have been included or excluded; what statistical relations have been assumed or derived; and what evidence supports these assumptions or derivations. All this information may not be included in the published paper, but it should be available to the reviewer. The overall aim of published reports should be to ensure transparency so that the importance and applicability of the methods can be clearly judged (see section 9)." |
| The choice of model used and the key parameters on which it is based are justified | If it's an RCT, single study ocst analysis (no modeling) | A justification for the choice of model and parameters used is not given. | A justification is given for the choice of model and key parameters used but it is not appropriate | A justification for the choice of model and key parameters used is given and is appropriate or sufficient for the question being asked. |
| Time horizon of costs and benefits is stated |  |  | Time horizon of costs and benefits is given but not appropriately justified | Time horizon of costs and benefits is given and appropriately justified. BMJ: "The time horizon should be long enough to capture all the differential effects of the options. It should often extend to the whole life of the treated individuals and even to future generations. If the time horizon is shortened for practical reasons this decision should be justified and an estimate made of any possible bias introduced. Justifying a short time horizon on the grounds of the duration of the available empirical evidence may be fallacious. If the relevant horizon for the decision is long term additional assumptions may need to be made.” |
| The discount rate(s) is stated | costs and benefits are not discounted | discount rate not stated |  | The discount rate is stated |
| The choice of rate(s) is justified | not applicable if discount rates aren't stated | the discount rate is not justified | a discount given for the discount rate is justified but that justification is not appropriate. If they just cite the authority (NICE or Neumann, for instance), just give a 1 | A justification for the choice of discount rate is given and is appropriate. BMJ: "At present most recommendations seem to vary between 3 and 6%, and a common rate in the literature is 5% per year. Certainly the analyst should use the government recommended rate, probably as the baseline value, and provide a sensitivity analysis with other discount rates. It is also helpful to provide the undiscounted data to allow the reader to recalculate the results using any discount rate...Most analysts argue that health benefits should be discounted at the same rate as costs in the baseline analysis, even if they are expressed in non-monetary units, such as life years or quality adjusted life years. A zero discount rate-or one lower than that used for costs-can be introduced in the sensitivity analysis. A lower rate is advocated so as not to penalize preventive programs and also because the results of some studies seem to suggest it...Given the current debates about discounting, the main emphasis should be on transparency in reporting the methods used." |
| An explanation is given if costs or benefits are not discounted | Costs and benefits are discounted | An explanation is not given for why costs and benefits are not discounted | An explanation is given for why costs and benefits are not discounted but is not sufficient | An explanation is given for why costs and benefits are not discounted and is sufficient |
| Details of statistical tests and confidence intervals are given for stochastic data | Not RCT (numbers are not coming from a trial study) | Details of statistical tests and confidence intervals are not given for stochastic data. |  | How the uncertainty ranges of model outcomes are derived (for example, from a probabilistic sensitivity analysis) is clearly stated. BMJ: "When stochastic data are reported details should be given of the statistical tests performed and the confidence intervals around the main variables." |
| The approach to sensitivity analysis is given | No sensitivity analysis | The approach to sensitivity analysis is not given. | The approach to sensitivity analysis is given but is not appropriate. | BMJ: "When a sensitivity analysis is performed details should be given of the approach used-for example, multivariate, uni-variate, threshold analysis...Simple sensitivity analysis (one way or multi-way), threshold analysis, analysis of extremes, and probabilistic sensitivity analysis may each be appropriate in particular circumstances." Paraphrased from BMJ: three types of uncertainty are recognized: uncertainty relating to observed data inputs, relating to extrapolation, relating to analytical methods. |
| The choice of variables for sensitivity analysis is justified | No sensitivity analysis | No justification is given for the choice of variables for sensitivity analysis. | Justification is given but is not appropriate or sufficient | BMJ: "justification given for the choice of variables for sensitivity analysis and the ranges over which they are varied." |
| The ranges over which the variables are varied are stated | No sensitivity analysis | The ranges over which the variables are varied are not stated | The ranges over which the variables are varied is stated but not appropriate or not stated clearly/fully | The ranges over which the variables are varied is stated and is appropriate. |
| Relevant alternatives are compared |  | No alternatives are compared. | Alternatives are compared, but are not relevant for the research question (whether due to dissimilarity in study methods or study settings) | BMJ: "Any comparisons with other health care interventions-for example, in terms of relative cost effectiveness-should be made only when close similarity in study methods and settings can be demonstrated...Beyond the individual study the reporting and interpretation of cost effectiveness ratios need to be handled with care. For example, authors often compare the cost effectiveness ratios generated in their own study with those for other interventions evaluated in previous studies in "league tables," where rankings are produced, ranging from the intervention with the lowest cost per". |
| Incremental analysis is reported | Not relevant to the original research question (i.e., league tables are not always the best decision tool) | Incremental analysis is not reported. |  | BMJ: "An incremental analysis--for example, incremental cost per life year gained--should be reported, comparing the relevant alternatives." |
| Major outcomes are presented in a disaggregated as well as aggregated form |  | Major outcomes are only presented in either disaggregated or aggregated form. | Either only aggregated or disaggregated outcomes are reported, or not every outcome has both. | BMJ: "The main emphasis in the reporting of study results should be on transparency. The main components of cost and benefit--for example, direct costs, indirect costs, life years gained, improvements in quality of life--should be reported in a disaggregated form before being combined in a single index or ratio...Reporting disaggregated data allows the reader to calculate other ratios that he or she sees fit." |
| The answer to the study question is given |  | The answer to the study question is given. | The answer to the study question is given but it is not appropriate or complete. | The answer to the study question is given and is appropriate based on the data. |
| Conclusions follow from the data reported |  | Conclusions from the data are not reported. | Conclusions are stated but they are not closely tied to the data. | Conclusions are stated and directly follow from the data. |
| Conclusions are accompanied by the appropriate caveats |  | No conclusions are given | Conclusions are just stated or stated with inappropriate caveats. | Conclusions are stated with appropriate caveats - i.e. qualifications or reservations |

| **Appendix Table 5: Additional Model Constructs Captured beyond BMJ Checklist Items*** | | | | | | | | | | |
| --- | --- | --- | --- | --- | --- | --- | --- | --- | --- | --- |
| Study | A probabilistic sensitivity analysis was conducted and the authors report some sort of interval (e.g. Confidence Interval, Uncertainty Interval) | Costs were based on primary sources (and not just copied from prior, similar studies). | Intervention and Healthcare Costs are Disaggregated | Formal Meta-Analysis Conducted | Systematic Literature Review Conducted | Model Validation Conducted | Population size based on real world population | Tornada diagram interpretable? | Explicit, isolated discussion of the assumptions | Explicitly acknowledged a peer-review set of guidelines |
| Naylor 2014^1^ | 0 | 0 | 0 | 0 | 0 | 0 | 0 | 1 | 0 | 0 |
| Rubio Terrés 2015^2^ | 0 | 1 | 0 | 0 | 0 | 0 | 0 | 0 | 0 | 0 |
| Ngeow 2015^3^ | 0 | 1 | 0 | 0 | 0 | 0 | 0 | 2 | 2 | 0 |
| Manchanda 2015^4^ | 0 | 2 | 0 | 0 | 0 | 0 | 2 | 2 | 0 | 0 |
| Barzi 2015^5^ | NR | 0 | 0 | 0 | 0 | 0 | 0 | NR | 2 | 0 |
| Snowsill 2015^6^ | NR | 2 | 2 | 0 | 0 | 0 | 0 | 2 | 2 | 0 |
| Gallego 2015^7^ | 2 | 1 | 0 | 0 | 0 | 0 | 2 | 2 | 0 | 0 |
| Bennette 2015^8^ | 2 | 0 | 0 | 0 | 0 | 0 | 0 | NR | 0 | 0 |
| Patel 2018^9^ | 0 | 2 | 0 | 0 | 0 | 0 | 0 | 2 | 0 | 0 |
| Johnson 2019^10^ | NR | 2 | 0 | 0 | 0 | 0 | 2 | 0 | 0 | 0 |
| Compagni 2013^11^ | 0 | 2 | 0 | 0 | 0 | 0 | 0 | 2 | 0 | 0 |
| Ademi 2019^12^ | 2 | 2 | 2 | 0 | 0 | 0 | 0 | NR | 0 | 0 |
| Gansen 2019^13^ | 0 | NA | 0 | 0 | 0 | 0 | 2 | NR | 0 | 0 |
| Catchpool 2019^14^ | 0 | 2 | 0 | 0 | 0 | 0 | 0 | 0 | 0 | 0 |
| Neusser 2019^15^ | 0 | 2 | 0 | 0 | 0 | 0 | 2 | NR | 0 | 0 |
| Asphaug 2019^16^ | 0 | 2 | 0 | 2 | 2 | 2 | 0 | NR | 0 | 0 |
| Hoskins 2019^17^ | 0 | 1 | 2 | 0 | 0 | 0 | 2 | NR | 0 | 0 |
| Vrijenhoek 2018^18^ | NR | 2 | 0 | 0 | 0 | 0 | 2 | NR | 0 | 0 |
| Kemp 2019^19^ | 0 | 0 | 0 | 0 | 0 | 0 | 0 | NR | 0 | 2 |
| Chen 2016^20^ | 0 | 2 | 2 | 0 | 0 | 0 | 2 | 2 | 0 | 0 |
| Nguyen 2017^21^ | 0 | 2 | 0 | 0 | 0 | 0 | 2 | 0 | 0 | 0 |
| Hayeems 2017^22^ | NR | 2 | 0 | 0 | 0 | 0 | 2 | NR | 0 | 0 |
| Eccleston 2017^23^ | 2 | 1 | 2 | 0 | 0 | 0 | 2 | NR | 2 | 0 |
| Li 2017^24^ | 2 | 0 | 0 | 0 | 0 | 0 | 0 | 2 | 0 | 0 |
| Kerr 2017^25^ | 0 | 0 | 2 | 0 | 0 | 0 | 0 | NR | 0 | 0 |
| Chen 2015^26^ | 0 | 0 | 0 | 0 | 0 | 0 | 0 | 0 | 0 | 0 |
| Crossland 2018^27^ | 2 | 2 | 0 | 2 | 2 | 0 | 2 | NR | 2 | 0 |
| Graaf 2017^28^ | 2 | 2 | 0 | 0 | 0 | 2 | 0 | NR | 0 | 0 |
| Manchanda 2017^29^ | 0 | 0 | 0 | 0 | 0 | 0 | 0 | NR | 0 | 0 |
| Manchanda 2018^30^ | 0 | 2 | 0 | 0 | 0 | 0 | 2 | NR | 0 | 0 |
| Severin 2015^31^ | 0 | 1 | 0 | 0 | 0 | 0 | 2 | 2 | 0 | 0 |
| Bonfanti 2016^32^ | 0 | 2 | 2 | 0 | 0 | 0 | 2 | NR | 2 | 0 |
| Goverde 2016^33^ | NR | 2 | 2 | 0 | 0 | 0 | 2 | 0 | 0 | 0 |
| Snowsill 2017^34^ | 0 | 2 | 0 | 2 | 2 | 0 | 2 | NR | 0 | 0 |
| Lázaro 2017^35^ | 0 | 2 | 2 | 0 | 0 | 0 | 2 | NR | 2 | 0 |
| Stark 2018^36^ | 2 | 2 | 2 | 0 | 0 | 0 | 2 | NR | 0 | 0 |
| Farnaes 2018^37^ | 0 | 2 | 0 | 0 | 0 | 0 | 2 | NR | 0 | 0 |
| McKay 2018^38^ | 0 | 2 | 0 | 2 | 2 | 0 | 0 | NR | 0 | 0 |
| Pelczarska 2018^39^ | 0 | 2 | 0 | 0 | 0 | 0 | 0 | NR | 0 | 0 |
| Lim 2018^40^ | 0 | 2 | 0 | 0 | 0 | 0 | 0 | 2 | 0 | 2 |
| Muram 2013^41^ | 0 | 2 | 0 | 0 | 0 | 0 | 0 | NR | 2 | 0 |
| Kwon 2019^42^ | 0 | 2 | 0 | 0 | 0 | 0 | 0 | NR | 0 | 0 |
| Muller 2019^43^ | 0 | 1 | 2 | 0 | 0 | 0 | 0 | NR | 2 | 0 |
| Schofield 2019^44^ | 0 | 2 | 0 | 0 | 0 | 0 | 2 | NR | 2 | 0 |
| Tuffaha 2018^45^ | 0 | 2 | 0 | 0 | 0 | 0 | 0 | NR | 0 | 0 |
| Zhang 2019^46^ | 2 | 2 | 2 | 0 | 0 | 0 | 2 | NR | 2 | 2 |
| Leenen 2016^47^ | NR | 2 | 2 | 0 | 0 | 0 | 2 | 0 | 0 | 0 |

**Assessment process was followed exactly as was applied for the 35 BMJ checklist items.*

| **Appendix Table 6A – BMJ Checklist Reference Numbers** | |
| --- | --- |
| **Item Number** | **BMJ Checklist Item** |
| 1 | The research question is stated |
| 2 | The economic importance of the research question is stated |
| 3 | The viewpoint(s) of the analysis are clearly stated and justified |
| 4 | The rationale for choosing the alternative programmes or interventions compared is stated |
| 5 | The alternatives being compared are clearly described |
| 6 | The form of economic evaluation used is stated |
| 7 | The choice of form of economic evaluation is justified in relation to the questions addressed |
| 8 | The source(s) of effectiveness estimates used are stated |
| 9 | Details of the design and results of effectiveness study are given (if based on a single study) |
| 10 | Details of the method of synthesis or meta-analysis of estimates are given (if based on an overview of a number of effectiveness studies) |
| 11 | The primary outcome measure(s) for the economic evaluation are clearly stated |
| 12 | Methods to value health states and other benefits are stated |
| 13 | Details of the subjects from whom valuations were obtained are given |
| 14 | Productivity changes (if included) are reported separately |
| 15 | The relevance of productivity changes to the study question is discussed |
| 16 | Quantities of resources are reported separately from their unit costs |
| 17 | Methods for the estimation of quantities and unit costs are described |
| 18 | Currency and price data are recorded (year, currency of costs, break into key components) |
| 19 | Details of currency of price adjustments for inflation or currency conversion are given |
| 20 | Details of any model used are given |
| 21 | The choice of model used and the key parameters on which it is based are justified |
| 22 | Time horizon of costs and benefits is stated |
| 23 | The discount rate(s) is stated |
| 24 | The choice of rate(s) is justified |
| 25 | An explanation is given if costs or benefits are not discounted |
| 26 | Details of statistical tests and confidence intervals are given for stochastic data |
| 27 | The approach to sensitivity analysis is given |
| 28 | The choice of variables for sensitivity analysis is justified |
| 29 | The ranges over which the variables are varied are stated |
| 30 | Relevant alternatives are compared |
| 31 | Incremental analysis is reported |
| 32 | Major outcomes are presented in a disaggregated as well as aggregated form |
| 33 | The answer to the study question is given |
| 34 | Conclusions follow from the data reported |
| 35 | Conclusions are accompanied by the appropriate caveats |

| **Appendix Table 6B: BMJ Checklist Assessment Across all Included Studies** | | | | | | | | | | | | | | | | | | | | | | | | | | | | | | | | | | | |
| --- | --- | --- | --- | --- | --- | --- | --- | --- | --- | --- | --- | --- | --- | --- | --- | --- | --- | --- | --- | --- | --- | --- | --- | --- | --- | --- | --- | --- | --- | --- | --- | --- | --- | --- | --- |
| Study (First Author, Year of Publication) | **BMJ Checklist Items** | | | | | | | | | | | | | | | | | | | | | | | | | | | | | | | | | | |
|  | 1 | 2 | 3 | 4 | 5 | 6 | 7 | 8 | 9 | 10 | 11 | 12 | 13 | 14 | 15 | 16 | 17 | 18 | 19 | 20 | 21 | 22 | 23 | 24 | 25 | 26 | 27 | 28 | 29 | 30 | 31 | 32 | 33 | 34 | 35 |
| Naylor 2014^1^ | 2 | 1 | 1 | 2 | 2 | 1 | 2 | 1 | NR | 0 | 2 | 0 | 0 | NR | NR | 0 | 1 | 1 | 0 | 2 | 2 | 2 | 2 | 0 | NR | 0 | 2 | 2 | 2 | 2 | 2 | 2 | 2 | 2 | 2 |
| Rubio Terrés 2015^2^ | 2 | 2 | 2 | 2 | 2 | 2 | 2 | 2 | 2 | 1 | 2 | 0 | NA | NR | NR | 0 | 1 | 1 | 0 | 2 | 2 | 2 | 2 | 0 | NR | 0 | 2 | 2 | 2 | 2 | 2 | 2 | 2 | 2 | 2 |
| Ngeow 2015^3^ | 2 | 0 | 1 | 1 | 2 | 1 | 2 | 2 | NR | 1 | 2 | 0 | 0 | 0 | 1 | 0 | 1 | 2 | 2 | 2 | 2 | 1 | 2 | 2 | NR | 0 | 2 | 2 | 2 | 2 | 2 | 2 | 2 | 2 | 2 |
| Manchanda 2015^4^ | 2 | 2 | 1 | 2 | 2 | 2 | 2 | 2 | 1 | NR | 2 | 2 | 2 | NR | NR | 2 | 2 | 2 | 2 | 2 | 2 | 2 | 2 | 2 | NR | 2 | 2 | 2 | 2 | 2 | 2 | 2 | 2 | 2 | 2 |
| Barzi 2015^5^ | 2 | 1 | 1 | 2 | 2 | 2 | 2 | 2 | NR | 0 | 2 | NR | NR | 0 | 1 | 0 | 1 | 0 | 0 | 2 | 2 | 2 | 2 | 2 | NR | 0 | 2 | 1 | 2 | 2 | 2 | 2 | 2 | 2 | 1 |
| Snowsill 2015^6^ | 2 | 2 | 2 | 2 | 2 | 2 | 2 | 2 | NR | 2 | 2 | 2 | 2 | NR | NR | 2 | 2 | 2 | 2 | 2 | 2 | 2 | 2 | 2 | NR | NR | 2 | 2 | 2 | 2 | 2 | 2 | 2 | 2 | 2 |
| Gallego 2015^7^ | 2 | 1 | 0 | 2 | 2 | 2 | 2 | 2 | NR | NR | 2 | 0 | 0 | NR | NR | 1 | 2 | 2 | 2 | 2 | 2 | 0 | 2 | 1 | NR | 0 | 2 | 2 | 2 | 2 | 2 | 2 | 2 | 2 | 2 |
| Bennette 2015^8^ | 2 | 1 | 1 | 2 | 2 | 2 | 2 | 2 | NR | NR | 2 | 0 | 0 | NR | NR | 1 | 1 | 2 | 0 | 1 | 2 | 2 | 2 | 0 | NR | 0 | 2 | 2 | 2 | 2 | 2 | 2 | 2 | 2 | 2 |
| Patel 2018^9^ | 2 | 2 | 1 | 2 | 2 | 2 | 2 | 2 | 1 | NR | 2 | 0 | 0 | NR | NR | 0 | 2 | 2 | 2 | 2 | 2 | 2 | 2 | 2 | NR | 0 | 2 | 2 | 2 | 2 | 2 | 2 | 2 | 2 | 2 |
| Johnson 2019^10^ | 2 | 2 | 2 | 2 | 2 | 2 | 2 | 2 | 2 | NR | 2 | 0 | 0 | NR | NR | 2 | 1 | 2 | 1 | 2 | 2 | 2 | 2 | 0 | NR | NR | 2 | 2 | 2 | 2 | 2 | 2 | 2 | 2 | 2 |
| Compagni 2013^11^ | 2 | 2 | 2 | 2 | 2 | 2 | 2 | 2 | NR | 1 | 2 | 0 | 1 | NR | NR | 2 | 2 | 1 | 0 | 2 | 2 | 2 | 2 | 2 | NR | 0 | 2 | 0 | 2 | 0 | 2 | 2 | 2 | 2 | 2 |
| Ademi 2019^12^ | 2 | 1 | 2 | 2 | 2 | 2 | 2 | 2 | 2 | NR | 2 | 0 | 0 | NR | NR | 0 | 1 | 2 | 2 | 2 | 2 | 2 | 2 | 1 | NR | 2 | 2 | 2 | 2 | 2 | 2 | 2 | 2 | 2 | 2 |
| Gansen 2019^13^ | 2 | 0 | 2 | 2 | 2 | 2 | 2 | 2 | NR | 2 | 2 | 2 | 2 | NR | NR | NA | NA | 2 | 2 | NA | NA | 2 | 2 | 0 | NR | NA | 2 | 2 | 2 | 2 | 2 | 2 | 2 | 2 | 2 |
| Catchpool 2019^14^ | 2 | 1 | 2 | 2 | 2 | 2 | 2 | 2 | 2 | NR | 2 | 2 | 2 | NR | NR | 0 | 2 | 2 | 2 | 2 | 2 | 2 | 2 | 2 | NR | 0 | 2 | 2 | 2 | 2 | 2 | 2 | 2 | 2 | 2 |
| Neusser 2019^15^ | 2 | 2 | 2 | 2 | 2 | NR | NR | 2 | 2 | NR | 2 | NR | NR | 0 | 0 | 2 | 2 | 2 | 2 | 2 | 2 | 2 | 2 | 0 | NR | 0 | 2 | 2 | 2 | 2 | 2 | 2 | 2 | 2 | 2 |
| Asphaug 2019^16^ | 2 | 1 | 1 | 2 | 2 | 2 | 2 | 2 | NR | 2 | 2 | 2 | 2 | 0 | 0 | 2 | 2 | 2 | 2 | 2 | 2 | 2 | 2 | 2 | NR | 2 | 2 | 2 | 2 | 2 | 2 | 2 | 2 | 2 | 2 |
| Hoskins 2019^17^ | 2 | 0 | 2 | 2 | 2 | 2 | 2 | NA | NR | 0 | 2 | 0 | 0 | NR | NR | 0 | 0 | 0 | 0 | 2 | 2 | 2 | 2 | 1 | NR | 0 | 2 | 2 | 2 | 2 | 2 | 2 | 2 | 2 | 1 |
| Vrijenhoek 2018^18^ | 2 | 2 | 2 | 2 | 2 | NR | NR | 2 | 2 | NR | 2 | NR | NR | NR | NR | 2 | 2 | 0 | 0 | NR | NR | NR | NR | NR | NR | NR | NR | NR | NR | 2 | NR | NR | 2 | 2 | 2 |
| Kemp 2019^19^ | 2 | 0 | 2 | 2 | 1 | 2 | 2 | NA | 2 | NR | 2 | NA | NA | NR | NR | NA | NA | NA | NA | NA | 1 | 2 | 2 | 2 | NR | 0 | 1 | NA | NA | 1 | 2 | 2 | 2 | 2 | 1 |
| Chen 2016^20^ | 2 | 2 | 2 | 2 | 2 | 2 | 2 | 2 | NR | 2 | 2 | NR | NR | NR | NR | 0 | NA | 2 | 2 | 2 | 2 | 1 | 2 | 0 | NR | 0 | 2 | 0 | 2 | 2 | 2 | 2 | 2 | 2 | 2 |
| Nguyen 2017^21^ | 2 | 2 | 0 | 2 | 2 | 2 | 2 | 2 | 2 | NR | 2 | NA | 1 | NR | NR | 2 | 2 | 2 | 2 | 2 | 2 | 2 | 2 | 0 | NR | 0 | 2 | 0 | 2 | 2 | 2 | 2 | 2 | 2 | 2 |
| Hayeems 2017^22^ | 2 | 2 | 1 | 2 | 1 | NR | NR | NR | 2 | NR | 2 | NR | 2 | NR | NR | 2 | 2 | 2 | 0 | 2 | NR | NR | NR | NR | NR | 2 | NR | NR | NR | 2 | NR | NR | 2 | 2 | 2 |
| Eccleston 2017^23^ | 2 | 0 | 2 | 2 | 2 | 2 | 2 | 2 | NR | 2 | 2 | 2 | 2 | NR | NR | 2 | NA | 2 | 0 | 2 | 2 | 2 | 2 | 2 | NR | 2 | 2 | 0 | 2 | 2 | 2 | 2 | 2 | 2 | 2 |
| Li 2017^24^ | 2 | 2 | 2 | 2 | 2 | 2 | 2 | 2 | NR | 2 | 2 | NA | NA | NR | NR | 0 | 1 | 2 | 2 | 2 | 2 | 2 | 2 | 0 | NR | 0 | 2 | 2 | 2 | 0 | 2 | 2 | 2 | 2 | 2 |
| Kerr 2017^25^ | 2 | 1 | 2 | 2 | 1 | 2 | 2 | 2 | NR | 1 | 2 | 0 | 0 | NR | NR | 0 | 0 | 2 | 0 | 2 | 2 | 1 | 2 | 2 | NR | 0 | 2 | 2 | 1 | 2 | 2 | 2 | 2 | 2 | 2 |
| Chen 2015^26^ | 2 | 2 | 2 | 2 | 2 | 2 | 2 | 2 | NR | 0 | 2 | 2 | 1 | NR | NR | 1 | 2 | 2 | 2 | 2 | 2 | 2 | 2 | 2 | NR | 0 | 2 | 2 | 2 | 2 | 2 | 2 | 2 | 2 | 2 |
| Crossland 2018^27^ | 2 | 1 | 2 | 2 | 2 | 2 | 2 | 2 | NR | 2 | 2 | 0 | 0 | NR | NR | 2 | 2 | 1 | 0 | 2 | 2 | 2 | 2 | 2 | NR | 0 | 2 | 1 | 2 | 0 | 2 | 2 | 2 | 2 | 2 |
| Graaf 2017^28^ | 2 | 0 | 2 | 2 | 2 | 2 | 2 | 2 | NR | 1 | 2 | 2 | 2 | NR | NR | 0 | 2 | 2 | 2 | 2 | 2 | 2 | 2 | 2 | NR | 1 | 2 | 2 | 2 | 2 | 2 | 2 | 2 | 2 | 2 |
| Manchanda 2017^29^ | 2 | 2 | 2 | 2 | 2 | 2 | 2 | 2 | NR | 0 | 2 | 0 | NA | NR | NR | 2 | 2 | 2 | 2 | NA | NA | 2 | 2 | 1 | NR | 0 | 2 | 0 | 2 | 2 | 2 | 2 | 2 | 2 | 1 |
| Manchanda 2018^30^ | 2 | 1 | 2 | 2 | 2 | 2 | 2 | 2 | NR | 2 | 2 | 2 | 0 | NR | NR | 2 | 2 | 2 | 2 | 2 | 2 | 2 | 2 | 2 | NR | 0 | 1 | 0 | 2 | 1 | 2 | 2 | 2 | 2 | 2 |
| Severin 2015^31^ | 2 | 1 | 2 | 2 | 2 | 2 | 2 | 2 | NR | 1 | 2 | NR | NR | NR | NR | 0 | 2 | 2 | 2 | 2 | 2 | 2 | 2 | 0 | NR | NR | 2 | 1 | 2 | 2 | 2 | 2 | 2 | 2 | 2 |
| Bonfanti 2016^32^ | 2 | 2 | 0 | 2 | 1 | 2 | 2 | 2 | 2 | NR | 2 | NR | NR | NR | NR | 1 | 2 | 2 | 0 | 1 | 2 | 2 | 1 | 0 | 0 | 0 | 0 | 0 | 0 | 2 | 0 | 2 | 2 | 2 | 2 |
| Goverde 2016^33^ | 2 | 0 | 0 | 2 | 2 | 2 | 2 | 2 | 2 | NR | 2 | NR | NR | NR | NR | 0 | 1 | 2 | 2 | 2 | 2 | 0 | 2 | 0 | NR | 0 | 1 | 1 | 2 | 2 | 2 | 2 | 2 | 2 | 2 |
| Snowsill 2017^34^ | 2 | 2 | 2 | 2 | 2 | 2 | 2 | 2 | NR | 2 | 2 | 2 | 2 | NR | NR | 2 | 2 | 2 | 2 | 2 | 2 | 2 | 2 | 2 | NR | NR | 2 | 2 | 2 | 2 | 2 | 2 | 2 | 2 | 2 |
| Lázaro 2017^35^ | 2 | 1 | 2 | 2 | 2 | 2 | 2 | 2 | NR | 0 | 2 | NA | 2 | 2 | 2 | 0 | 2 | 1 | 0 | 2 | 2 | 2 | 2 | 0 | NR | 0 | 2 | 0 | 2 | 2 | 2 | 2 | 2 | 2 | 1 |
| Stark 2018^36^ | 2 | 2 | 2 | 1 | 2 | 2 | 2 | 2 | 2 | NR | 2 | 1 | 2 | NR | NR | 1 | 1 | 1 | 2 | 0 | 1 | 1 | 0 | 0 | 0 | 2 | 1 | NR | NR | 1 | 2 | 2 | 2 | 2 | 2 |
| Farnaes 2018^37^ | 2 | 2 | 2 | 2 | 2 | NR | NR | 2 | 2 | NR | 2 | NR | NR | NR | NR | 0 | 0 | 0 | 0 | 2 | 2 | NR | NR | NR | NR | 2 | NR | NR | NR | NR | NR | NR | 2 | 2 | 2 |
| McKay 2018^38^ | 2 | 2 | 2 | 2 | 2 | 2 | 2 | 2 | NR | 2 | 2 | NA | NA | NR | NR | 1 | 2 | 2 | 1 | 2 | 2 | 2 | 2 | 2 | NR | 0 | 1 | 2 | 2 | 2 | 2 | 2 | 2 | 2 | 2 |
| Pelczarska 2018^39^ | 2 | 1 | 1 | 1 | 1 | 2 | 2 | 2 | NA | NA | 2 | 1 | 1 | NR | NR | 0 | 1 | 2 | 1 | 2 | 2 | 2 | 2 | 2 | NR | 0 | 2 | 1 | 2 | 2 | 2 | 2 | 2 | 2 | 2 |
| Lim 2018^40^ | 2 | 2 | 2 | 2 | 2 | 2 | 2 | 2 | NR | 1 | 2 | 2 | 2 | NR | NR | 1 | 2 | 2 | 2 | 2 | 2 | 2 | 2 | 2 | NR | 0 | 2 | 2 | 2 | 2 | 2 | 2 | 2 | 2 | 2 |
| Muram 2013^41^ | 2 | 2 | 2 | 2 | 2 | NR | 2 | 2 | 2 | NR | 2 | NR | NR | NR | NR | 0 | 2 | 2 | 2 | 2 | 2 | 1 | 2 | 0 | NR | 0 | NR | NR | NR | 0 | 2 | 2 | 2 | 2 | 2 |
| Kwon 2019^42^ | 2 | 0 | 1 | 2 | 2 | 2 | 2 | 2 | NR | NR | 2 | NA | NA | NR | NR | 0 | 0 | 2 | 0 | 2 | 2 | 2 | 2 | 2 | NR | 0 | 1 | 2 | 2 | 0 | 2 | 2 | 2 | 2 | 2 |
| Muller 2019^43^ | 2 | 2 | 2 | 2 | 2 | 1 | 2 | 2 | NR | 1 | 2 | 2 | 1 | NR | NR | 1 | 2 | 1 | 0 | 2 | 2 | 2 | 2 | 2 | NR | 0 | 1 | 0 | 0 | 2 | 2 | 2 | 2 | 2 | 2 |
| Schofield 2019^44^ | 2 | 2 | 0 | 2 | 2 | 2 | 2 | 2 | 2 | NR | 1 | 2 | 2 | NR | NR | 1 | 2 | 2 | 2 | 1 | 1 | 2 | 2 | 2 | NR | 0 | 2 | 2 | 1 | 2 | 2 | 2 | 2 | 2 | 1 |
| Tuffaha 2018^45^ | 2 | 2 | 2 | 2 | 2 | 2 | 2 | 2 | NR | NR | 2 | NA | NA | NR | NR | 2 | 2 | 2 | NR | 2 | 2 | 2 | 2 | 0 | NR | 0 | 2 | 0 | 0 | 2 | 2 | 2 | 2 | 2 | 2 |
| Zhang 2019^46^ | 2 | 2 | 2 | 2 | 2 | 2 | 2 | 2 | NR | 2 | 2 | NA | 2 | NR | NR | 0 | 1 | 2 | 0 | 2 | 2 | 2 | 2 | 0 | NR | 2 | 2 | 2 | 2 | 2 | 2 | 2 | 2 | 2 | 2 |
| Leenen 2016^47^ | 2 | 0 | 1 | 2 | 2 | 2 | 2 | 2 | 2 | 1 | 2 | NR | NR | NR | NR | 0 | NA | 2 | 2 | 1 | 2 | 1 | 2 | 0 | NR | 0 | 1 | 0 | 2 | 2 | 2 | | 2 | 2 | 2 |

**Appendix 7: Study-Specific BMJ Checklist Values**

| **Appendix Table 7: BMJ Checklist Values across all included studies** | | | | | | |
| --- | --- | --- | --- | --- | --- | --- |
| Study (first author, year of publication) | Total 2s | Total 1s | Total 0s | Total NRs | Total NAs | Overall Value*  (from highest to lowest) |
| Snowsill 2015 | 38 | 0 | 4 | 7 | 0 | 90% |
| Snowsill 2017 | 39 | 0 | 5 | 5 | 0 | 89% |
| Hayeems 2017 | 15 | 2 | 1 | 17 | 0 | 89% |
| Manchanda 2015 | 37 | 3 | 4 | 5 | 0 | 88% |
| Farnaes 2018 | 14 | 0 | 2 | 19 | 0 | 88% |
| Bonfanti 2016 | 25 | 0 | 4 | 6 | 0 | 86% |
| Zhang 2019 | 34 | 1 | 6 | 4 | 2 | 84% |
| Asphaug 2019 | 38 | 2 | 7 | 2 | 0 | 83% |
| Vrijenhoek 2018 | 20 | 1 | 4 | 10 | 0 | 82% |
| Ademi 2015 | 35 | 3 | 7 | 4 | 0 | 81% |
| Catchpool 2019 | 36 | 1 | 8 | 4 | 0 | 81% |
| Graaff 2017 | 34 | 2 | 8 | 4 | 0 | 80% |
| Lim 2018 | 35 | 2 | 8 | 4 | 0 | 80% |
| Eccleston 2017 | 33 | 1 | 8 | 4 | 1 | 80% |
| McKay 2018 | 33 | 3 | 7 | 4 | 2 | 80% |
| Johnson 2019 | 32 | 2 | 8 | 7 | 0 | 79% |
| Gansen 2019 | 27 | 0 | 7 | 4 | 11 | 79% |
| Severin 2015 | 31 | 4 | 7 | 7 | 0 | 79% |
| Neusser 2019 | 14 | 0 | 4 | 17 | 0 | 78% |
| Tuffaha 2018 | 30 | 2 | 9 | 6 | 2 | 76% |
| Compagni 2013 | 32 | 3 | 9 | 5 | 0 | 76% |
| Chen 2015 | 33 | 2 | 10 | 4 | 0 | 76% |
| Manchanda 2018 | 30 | 6 | 8 | 5 | 0 | 75% |
| Patel 2018 | 32 | 2 | 10 | 5 | 0 | 75% |
| Rubio-Terrés 2015 | 30 | 4 | 10 | 4 | 1 | 73% |
| Crossland 2018 | 31 | 3 | 11 | 4 | 0 | 72% |
| Nguyen 2017 | 31 | 1 | 12 | 4 | 1 | 72% |
| Scholfield 2019 | 27 | 7 | 9 | 6 | 0 | 71% |
| Chen 2016 | 28 | 1 | 12 | 6 | 2 | 70% |
| Bennette 2015 | 28 | 6 | 10 | 5 | 0 | 70% |
| Ngeow 2015 | 28 | 8 | 10 | 3 | 0 | 70% |
| Lázaro 2017 | 29 | 4 | 12 | 2 | 2 | 69% |
| Gallego 2015 | 28 | 5 | 11 | 5 | 0 | 69% |
| Muller 2019 | 26 | 8 | 10 | 4 | 0 | 68% |
| Leenen 2016 | 24 | 6 | 10 | 8 | 1 | 68% |
| Manchanda 2017 | 26 | 2 | 12 | 4 | 5 | 68% |
| Stark 2018 | 25 | 9 | 10 | 5 | 0 | 67% |
| Li 2017 | 28 | 2 | 13 | 4 | 2 | 67% |
| Goverde 2016 | 25 | 3 | 12 | 9 | 0 | 66% |
| Kwon 2019 | 26 | 3 | 13 | 5 | 2 | 65% |
| Kemp 2019 | 19 | 5 | 9 | 4 | 12 | 65% |
| Pelczarska 2018 | 23 | 9 | 12 | 3 | 2 | 63% |
| Barzi 2015 | 24 | 6 | 13 | 6 | 0 | 63% |
| Muram 2013 | 17 | 4 | 9 | 5 | 0 | 63% |
| Hoskins 2019 | 23 | 4 | 14 | 4 | 4 | 61% |
| Kerr 2017 | 22 | 6 | 16 | 4 | 0 | 57% |
| Naylor 2014 | 20 | 9 | 16 | 4 | 0 | 54% |

*Overall quality values were calculated for each question by summing the 1s and 2s each article received across all studies then dividing that sum by the number of items for which 0s, 1s, and 2s were possible (items which were not coded NR or NA).

**Appendix 8: The Use of Scenario Analyses in Familial Hypercholesterolemia Studies**

For the sake of describing how scenario analyses can be best utilized in economic evaluations of genetic screening, we provide a detailed description of how such analyses were used across studies in review which focused on Familia Hypercholesterolemia (FH).

The value of screening for FH depends in part on how well people adhere to prophylactic treatment. Lipid modification treatment adherence among FH patients is likely imperfect and will vary across contexts. However, two studies explicitly assumed adherence would be 100% and one study made no explicit commentary, presumably assuming an adherence of 100%. An important improvement, McKay and colleagues performed a scenario analysis in which 15% of those on therapy would discontinue at 10 years and Chen and colleagues performed and analysis to estimate the impact a therapy adherence program would have on the cost-effectiveness of FH screening. Both McKay and Chen’s analysis determined that the cost-effectiveness of FH screening was highly sensitive to adherence, revealing the critical importance of conducting their respective sensitivity analysis and therefore the methodological limitations of the other FH studies. The inclusion of cascade testing of family members is also an important target of scenario analysis, as cascade testing dramatically increases the overall value of a screening program. Evaluating cascade testing using scenario analysis allows direct comparison of the value with or without the more cost-effective downstream implications for family members (who may or may not be covered within the same health plan). In McKay and colleagues’ analysis, four strategies are considered without cascade testing, and then three of those four strategies are considered with reverse cascade testing to determine the added efficacy of cascade testing.
